# Respiratory syncytial virus vaccination strategies for older Canadian adults: a cost-utility analysis

**DOI:** 10.1101/2024.03.20.24304630

**Authors:** Ashleigh R. Tuite, Alison E. Simmons, Monica Rudd, Alexandra Cernat, Gebremedhin B. Gebretekle, Man Wah Yeung, April Killikelly, Winnie Siu, Sarah A. Buchan, Nicholas Brousseau, Matthew Tunis

## Abstract

**Background:** Vaccines against respiratory syncytial virus (RSV) have the potential to reduce disease burden and costs in Canadians, but the cost-effectiveness of RSV vaccination programs for older adults is unknown. We evaluated the cost-effectiveness of different adult age cutoffs for RSV vaccination programs, with or without a focus on people with higher disease risk due to chronic medical conditions (CMCs).

**Methods:** We developed a static individual-based model of medically-attended RSV disease to evaluate the cost-utility of alternate age-, medical risk-, and age-plus medical risk-based vaccination policies. The model followed a multi-age cohort of 100,000 people aged 50 years and older over a three-year period. Vaccine characteristics were based on RSV vaccines authorized in Canada as of March 2024. We calculated incremental cost-effectiveness ratios (ICERs) in 2023 Canadian dollars per quality-adjust life year (QALY) from the health system and societal perspectives, discounted at 1.5%.

**Results:** Although all vaccination strategies averted medically-attended RSV disease, strategies focused on adults with CMCs were more likely to be cost-effective than age-based strategies. A program focused on vaccinating adults aged 70 years and older with one or more CMCs was optimal for a cost-effectiveness threshold of $50,000 per QALY. Results were sensitive to assumptions about vaccine price, but approaches based on medical risk remained optimal compared to age-based strategies even when vaccine prices were low. Findings were robust to a range of alternate assumptions.

**Interpretation:** Based on available data, RSV vaccination programs in some groups of older Canadians with underlying medical conditions are expected to be cost-effective.

## INTRODUCTION

Respiratory syncytial virus (RSV) infections cause a substantial burden of disease, particularly at the extremes of the age spectrum, with rates of medically-attended RSV highest in infants and older adults (1–4). Among adults, incidence of medically-attended RSV increases with age (5, 6). The presence of underlying medical conditions, such as chronic obstructive pulmonary disease, congestive heart failure, diabetes, asthma, and immunocompromise, is associated with more severe RSV disease in adults (7, 8).

The landscape of RSV disease prevention has evolved markedly in the past several years, with multiple immunization products now authorized for infants and adults (9). As of March 2024, there are two RSV vaccines approved for use in Canada in adults aged 60 years and older (Arexvy and Abrysvo) (9) and another under review (mRNA-1345) (10).

RSV vaccines have the potential to reduce healthcare and related costs in older Canadians but the introduction of publicly-funded vaccination programs may be costly. Based on available epidemiological data, not all older adults derive equal benefit from vaccination; those who are younger and without underlying medical conditions are expected to have a lower risk of severe RSV disease and may benefit less from vaccination.

Cost-effectivness analysis can be used to quantify the costs and benefits of potential RSV vaccination programs. To date, most economic analyses of RSV vaccination programs in older adults have focused on cost-effectiveness in adults age 60 or 65 years and older (11). Given the elevated risk of RSV disease with increasing age and in people with underlying medical conditions, we sought to evaluate the optimal use of RSV vaccines in the Canadian population. Specifically, we evaluated the cost-effectiveness of different age cutoffs for programs that were either focused on people at high risk of RSV disease or offered to the entire age group regardless of medical risk status.

## METHODS

### Model overview

We conducted a model-based cost-utility analysis of RSV vaccination programs in the Canadian population aged 50 years and older. We developed a static individual-based model of medically-attended RSV disease to explore the impact of alternate: (i) age-, (ii) medical risk-, and (iii) age-plus medical risk-based vaccination policies on RSV-associated outcomes. Costs were in 2023 Canadian dollars and, where necessary, were adjusted using the Canadian Consumer Price Index (12). A discount rate of 1.5% was used for costs and outcomes and cost-effectiveness was assessed from both the health system and societal perspectives (13). The model was constructed and analyzed using R (14). Full model details are provided in the **Supplementary Material**.

The model followed a multi-age closed population cohort of 100,000 people over a three-year period that included three full RSV seasons, with the age group distribution based on projections of the Canadian population aged 50 years and older (15). Individuals were further characterized by the presence or absence of one or more chronic medical conditions (CMCs) (16). Model start time occurred in September, at the onset of a new RSV season and used monthly time steps. A portion of the population was vaccinated in the first two months of model entry with vaccination coverage based on influenza vaccine uptake, which varied by age and CMC status (17). We included risks of solicited severe local and severe systemic adverse events following immunization (AEFI). RSV infection could occur at any month during the three-year period and was assumed to follow seasonal trends, with peak activity occurring from January to March. A three-year period was used based on currently available data showing that vaccine protection wanes over time but lasts for at least two RSV seasons (18–20). The three-year period allowed for investigation of vaccine protection that potentially lasts through three RSV seasons in scenario analysis.

We modelled medically-attended RSV disease only, with individuals requiring one of the following levels of care: outpatient (healthcare provider visit or emergency department (ED) visit), or inpatient (hospitalization, with or without intensive care unit (ICU) admission) (**Figure *1***). Vaccination reduced the risk of these outcomes. Although multiple RSV infections are possible within a season and in subsequent seasons (21), for simplicity, we assumed a maximum of one medically-attended RSV infection per person over the model time horizon.

**Figure 1.**
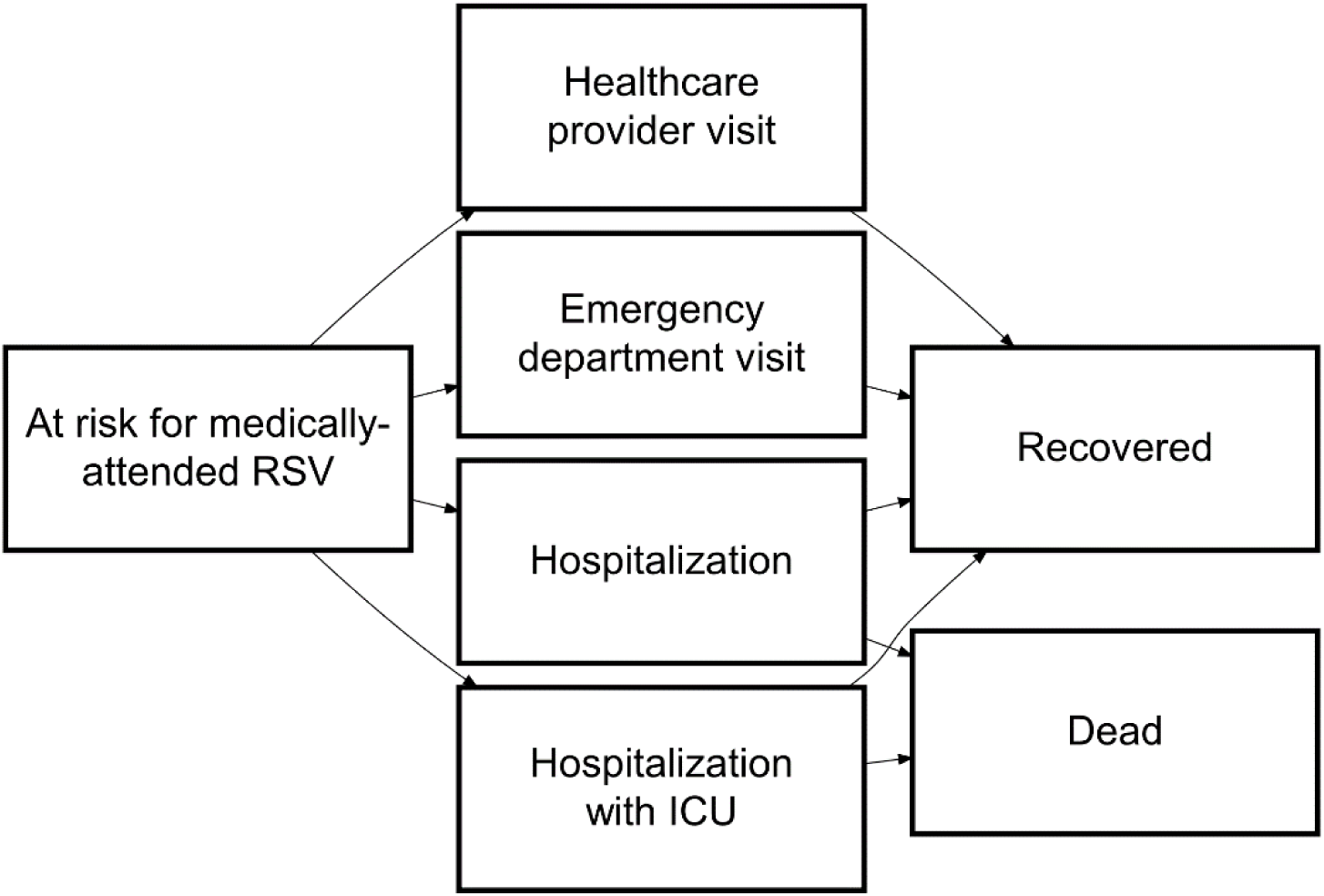
Health states included in the model associated with medically-attended RSV. Arrows indicate possible transitions between health states. Individuals can transition to death from any of the other health states due to background mortality (arrows not shown). Risk of experiencing any medically-attended RSV outcome varies by age, chronic medical condition status, and vaccination status.

Model parameters describing RSV epidemiology, vaccine characteristics, costs, and health utilities were obtained from published studies and available data, when possible, and by assumption or expert opinion otherwise (**Table 1**). Canadian data were used preferentially and we used age- and CMC-status specific estimates, when available. Ranges in **Table 1** indicate parameters that were drawn from distributions for the analysis, with beta distributions used for probabilities and utilities, and gamma distributions used for costs.

**Table 1.** Model input parameters.

| Parameter | Base | Range | Reference |
| --- | --- | --- | --- |
| Population distribution (%) |  |  |  |
| 50-59 years | 31.7 | -- | Statistics Canada (15) |
| 60-64 years | 17.5 | -- |  |
| 65-69 years | 15.8 | -- |  |
| 70-74 years | 12.8 | -- |  |
| 75-79 years | 9.9 | -- |  |
| 80+ years | 12.2 | -- |  |
| % of population with one or more CMCs |  |  |  |
| 50-59 years | 38.4 | -- | Statistics Canada (16) |
| 60-79 years | 60.1 | -- |  |
| 80+ years | 72.1 | -- |  |
| Monthly % of annual RSV cases |  |  |  |
| September | 1.2 | -- | Respiratory Virus Detection Surveillance System (average of 9 seasons, 2010-2011 to 2018-2019) (22) |
| October | 1.9 | -- |  |
| November | 5.5 | -- |  |
| December | 14.2 | -- |  |
| January | 17.6 | -- |  |
| February | 21.1 | -- |  |
| March | 17.0 | -- |  |
| April | 11.0 | -- |  |
| May | 5.6 | -- |  |
| June | 2.6 | -- |  |
| July | 1.3 | -- |  |
| August | 1.1 | -- |  |
| Odds ratio for medically-attended outpatient care in adults with one or more CMCs |  |  |  |
| All ages | 1.1 | -- | Shi et al. 2022 (7) |
| % of patients requiring hospitalization with one or more CMCs |  |  |  |
| All ages | 98.2 | -- | EISherif et al. 2023 (23) |
| Under-detection factor for medically-attended RSV in adults |  |  |  |
| All ages | 1.5 | 1-2 | McLaughlin et al. 2022 (8) |
| Annual incidence of medically-attended RSV requiring outpatient healthcare provider visit per 100,000 population (unadjusted for under-detection) |  |  |  |
| 50-59 years | 261.9 | 186.5-337.3 | EISherif et al. 2023 (23);<br>McLaughlin et al. 2022 (8);<br>Respiratory Virus Detection Surveillance System (22) |
| 60-69 years | 604.1 | 472.8-707.8 |  |
| 70-79 years | 780.0 | 625.1-934.1 |  |
| 80+ years | 2487.1 | 2097.1-2877.2 |  |
| Annual incidence of medically-attended RSV requiring emergency department visit per 100,000 population (unadjusted for under-detection) |  |  |  |
| 50-59 years | 16.8 | 12.0-21.6 |  |
| 60-69 years | 43.3 | 33.9-50.7 | ElSherif et al. 2023 (23);<br>McLaughlin et al. 2022 (8);<br>Respiratory Virus Detection<br>Surveillance System (22) |
| 70-79 years | 68.5 | 54.9-82.0 |  |
| 80+ years | 218.3 | 184.1-252.5 |  |
| Annual incidence of RSV-attributable hospitalization per 100,000 population (unadjusted for under-detection) |  |  |  |
| 50-59 years | 15.1 | 10.8-19.5 | ElSherif et al. 2023 (23);<br>Respiratory Virus Detection<br>Surveillance System (22) |
| 60-69 years | 47.5 | 37.2-55.7 |  |
| 70-79 years | 96.4 | 77.3-115.4 |  |
| 80+ years | 307.4 | 259.2-355.6 |  |
| % of patients hospitalized with RSV requiring ICU admission |  |  |  |
| All ages | 13.7 | 10.2-17.9 | ElSherif et al. 2023 (23) |
| Medically-attended outpatient RSV disease duration (days) |  |  |  |
| 50-64 years | 7 | 6-9 | Gessner 2000 (49);<br>assumption |
| 65+ years | 15 | 13-18 | Falsey et al. 2005 (50) |
| % of patients with medically attended RSV prescribed an antimicrobial |  |  |  |
| All ages | 50 | 14-89 | Bernardo et al. 2019 (51);<br>ElSherif et al. 2023 (23) |
| Length of stay in hospital (days) |  |  |  |
| 50-64 years | 6 | 3-13 | Chen et al. 2024 (52) |
| 65-74 years | 6 | 4-13 |  |
| 75+ years | 7 | 4-13 |  |
| Length of stay in ICU (days) |  |  |  |
| All ages | 5 | 3-9 | ElSherif et al. 2023 (23) |
| RSV mortality per hospitalization (%) |  |  |  |
| 50-64 years | 7.2 | 5.4-9.5 | Chen et al. 2024 (52) |
| 65-74 years | 6.6 | 5.2-8.4 |  |
| 75+ years | 10.1 | 9.0-11.3 |  |
| All-cause mortality rate (per year, per 1,000 population) |  |  |  |
| All ages | Age-specific rates | -- | Statistics Canada (53) |
| Immunization coverage (%), with chronic medical conditions |  |  |  |
| 50-59 years | 58.6 | -- | Seasonal Influenza<br>Vaccination Coverage<br>Survey, 2022-2023 (17) |
| 60-64 years | 59.9 | -- |  |
| 65-69 years | 65.2 | -- |  |
| 70-79 years | 82.7 | -- |  |
| 80+ years | 83.4 | -- |  |
| Immunization coverage (%), without chronic medical conditions |  |  |  |
| 50-59 years | 36.7 | -- | Seasonal Influenza<br>Vaccination Coverage<br>Survey 2022-2023 (17) |
| 60-64 years | 49.4 | -- |  |
| 65-69 years | 61.1 | -- |  |
| 70-79 years | 74.9 | -- |  |
| 80+ years | 74.8 | -- |  |
| Time to receive vaccine (hours), including travel and waiting time |  |  |  |
| All ages | 2 | 1.5-2.5 | Canada Health Infoway (54); Ray et al. 2015 (55) |
| Vaccine effectiveness (%), Arexvy (GSK) |  |  |  |
| Outpatient RSV – season 1 (7 mo follow-up) | 82.6 | -- | Friedland 2023 (19); Ison et al. 2024 (20); assumption for season 3 |
| Outpatient RSV – season 2 (6 mo follow-up) | 56.1 | -- |  |
| Outpatient RSV – season 3 | 18.7 | -- |  |
| Hospitalized RSV – season 1 (7 mo follow-up) | 94.1 | -- |  |
| Hospitalized RSV – season 2 (6 mo follow-up) | 64.2 | -- |  |
| Hospitalized RSV – season 3 | 21.4 | -- |  |
| Vaccine effectiveness (%), Abrysvo (Pfizer) |  |  |  |
| Outpatient RSV – season 1 (7 mo follow-up) | 65.1 | -- | Gurtman 2023 (18); assumption for season 3 |
| Outpatient RSV – season 2 (4 mo follow-up) | 48.9 | -- |  |
| Outpatient RSV – season 3 | 16.3 | -- |  |
| Hospitalized RSV – season 1 (7 mo follow-up) | 88.9 | -- |  |
| Hospitalized RSV – season 2 (4 mo follow-up) | 78.6 | -- |  |
| Hospitalized RSV – season 3 | 26.2 | -- |  |
| Vaccine wastage rate (%) |  |  |  |
| All ages | 5 | -- | WHO, 2019 (56) |
| Adverse events following immunization (%) |  |  |  |
| Severe local adverse event | 0.51 | 0.16-1.84 | Melgar et al. 2023 (24) |
| Severe systemic adverse event | 0.57 | 0.10-2.35 |  |
| Duration of adverse event following immunization (days) |  |  |  |
| Severe local adverse event | 1 | 1-4 | Papi et al. 2023 (57); Walsh et al. 2023 (58); Lee et al. 2009 (30) |
| Severe systemic adverse event | 2 | 1-4 | Papi et al. 2023 (57); Walsh et al. 2023 (58); Lee et al. 2009 (30) |
| Cost of vaccine administration per dose (\$) | | | |
| All ages | 18 | 13-22 | O'Reilly et al. 2017 (27) |
| Immunization cost per dose (\$) | | | |
| GSK | 230 | 100-230 | Robertson, 2023 (28) |
| Pfizer | 230 | 100-230 |  |
| Attributable costs per person hospitalized with RSV (\$) | | | |
| Hospitalization (6 months) | 32,228 | 31,622-32,836 | Mac et al. 2023 (25) |
| Hospitalization, die in hospital | 27,534 | 22,027-33,041* |  |
| Costs per person with RSV treated in the outpatient setting (\$) | | | |
| Healthcare provider visit | 62 | 48-82 | Sander et al. 2010 (26);<br>CIHI (29); Alliance for<br>Healthier Communities (59) |
| ED visit | 340 | 302-509 |  |
| Direct medical costs for severe local adverse event following vaccination (\$) | | | |
| <65 years | 62 | 48-82 | Sander et al. 2010 (26);<br>CIHI (29); Lee et al. 2009<br>(30) |
| 65+ years | 63 | 49-83 |  |
| Direct medical costs for severe systemic adverse event following vaccination (\$) | | | |
| <65 years | 62 | 48-82 | Sander et al. 2010 (26);<br>CIHI (29); Lee et al. 2009<br>(30) |
| 65+ years | 66 | 51-87 |  |
| Transportation costs (\$) | | | |
| Cost of two-way travel to<br>vaccination or outpatient care (out<br>of pocket) | 14 | 11-16* | Canada Health Infoway<br>(39, 54) |
| Cost of travel to inpatient care | 417 | 210-623 | NACI (39) |
| Co-pay per prescription (\$) | | | |
| 50-64 years | 10 | 5-12 | University of Western<br>Ontario (60) |
| 65+ years | 10 | 7-34 | The Commonwealth Fund<br>(61); assumption |
| Medication costs (\$) | | | |
| Over-the-counter medication<br>costs for medically-attended RSV<br>case | 12 | 3-20 | Federici et al. 2018 (62) |
| Prescription medication costs for<br>medically-attended RSV case<br>(<65 years) | 14 | 6-22 | Federici et al. 2018 (62) |
| Prescription medication costs for<br>severe local adverse event<br>following vaccination (<65 years) | 1.05 | 0.79-1.31 | Lee et al. 2009 (30) |
| Prescription medication costs for<br>severe systemic adverse event<br>following vaccination (<65 years) | 4.19 | 3.14-5.24 | Lee et al. 2009 (30) |
| Caregiver workdays lost |  |  |  |
| % reduction in productivity | 33 | -- | Keita Fakeye et al. 2023<br>(63) |
| Labour force participation (%) |  |  |  |
| 50-59 years | 82.6 | -- | Statistics Canada (64) |
| 60-64 years | 58.5 | -- |  |
| 65-69 years | 29.3 | -- |  |
| 70+ years | 8.2 | -- |  |
| Caregiver | 88.8 | -- |  |
| Average employment income (\$) | | | |
| 50-59 years | 65,466 | -- | Statistics Canada (65) |
| 60-64 years | 62,674 | -- |  |
| 70 years and over | 27,721 | -- |  |
| Caregiver (age 25-54 years) | 68,372 | -- |  |
| Background health utility |  |  |  |
| 50-59 years | 0.848 | -- | Yan et al. 2023 (31) |
| 60-64 years | 0.839 | -- |  |
| 65-74 years | 0.867 | -- |  |
| 75+ years | 0.861 | -- |  |
| QALY loss, outpatient, with or without ED visit |  |  |  |
| All ages | 0.0056 | 0.0037 – 0.0075 | Herring et al. 2022 (33);<br>Mao et al. 2022 (34);<br>Zeevat et al. 2022 (35);<br>Meijboom et al. 2013 (32) |
| QALY loss, hospitalization |  |  |  |
| All ages | 0.020 | 0.017-0.030 | Herring et al. 2022 (33);<br>Mao et al. 2022 (34);<br>Zeevat et al. 2022 (35);<br>Meijboom et al. 2013 (32) |
| QALY loss, death |  |  |  |
| 50-59 years | 20.26 | -- | Yan et al. 2023 (31);<br>Statistics Canada (53, 66) |
| 60-64 years | 16.74 | -- |  |
| 65-69 years | 14.29 | -- |  |
| 70-74 years | 11.75 | -- |  |
| 75-79 years | 9.38 | -- |  |
| 80+ years | 5.84 | -- |  |
| QALY loss, adverse event following vaccination |  |  |  |
| Serious local adverse event | 0.0003 | 0.0002-0.0004 | Prosser et al. 2023 (36);<br>assumption |
| Serious systemic adverse event | 0.0004 | 0.0003-0.0005 |  |

\*Range defined as $\pm 20\%$ of the base value

The proportion of annual cases occurring each month was estimated from reported RSV tests and positive detections for all of Canada (22). Incidence of hospitalized RSV and risk of ICU admission were obtained from a Canadian study (23). Outpatient incidence of medically-attended RSV disease was estimated by applying rate ratios of healthcare provider or ED visits to hospitalized cases from a meta-analysis (8) to Canadian hospitalization rates. We accounted for the association of CMCs with increased risk of medically-attended RSV (4, 7) by adjusting age-specific outpatient and inpatient incidence estimates to be consistent with the reported fractions of people receiving outpatient care or hospitalized with RSV with at least one CMC (7, 23). We adjusted all RSV outcome estimates by an under-detection ratio of 1.5 in the base case analysis (8).

We used data for the two RSV vaccines (Arexvy (GSK), and Abrysvo (Pfizer)) currently authorized for use in Canada to estimate vaccine effectiveness (VE) and rates of AEFIs (18–20, 24). Data on average VE and average duration of follow-up for each season were used to generate step functions, with protection assumed to wane linearly between seasons. We used a cubic polynomial regression model to obtain smoothed estimates of VE over 36 months (**Supplementary Figure 1**); in the absence of data for season 3, we assumed that VE reached one-third of season 2 VE by the end of the season (i.e., month 36) to model waning past the end of RCT data. We assumed that VE did not vary by age or CMC status. Since RCT data were only available for up to two RSV seasons, in our base case analysis we conservatively assumed that VE in season 3 was 0. We modelled VE extending through to season 3 in a scenario analysis.

Costs of inpatient RSV were based on attributable costs derived from a retrospective population-based cohort study in Ontario, Canada (25). Costs for outpatient cases were based on estimates for influenza (26). Vaccination costs included administration costs and Canadian list prices (27, 28). Direct costs for AEFIs included a healthcare provider visit and treatment costs (26, 29, 30). Costs for the societal perspective included: patient productivity loss due to AEFIs and RSV-attributable illness and death; caregiver productivity loss; and out-of-pocket medical costs.

Age-specific utilities for the general population were based on EQ-5D-5L index scores for the Canadian population (31). QALY losses associated with the modelled health outcomes were derived from published studies and assumption (32–36).

### Vaccination strategies

We evaluated a combination of age-only, medical risk-only, and age plus medical risk-based single dose vaccination strategies (**Table 2**). For age-based strategies, all people aged greater than or equal to the specified age cutoff (i.e., 60, 65, 70, 75, or 80 years and older) were eligible to receive the vaccine. For medical risk-based strategies, only people aged greater than or equal to the specified age cutoff who also had one or more CMCs were eligible to receive the vaccine. For age-plus medical risk-based strategies, people were eligible to receive the vaccine if they met an age requirement, or a lower age requirement if they had at least one CMC. For age-plus medical risk-based strategies we evaluated a lower age bound for people with CMC of either 50 years or 60 years. Although the vaccines are currently authorized for use in adults aged 60 and older, we considered a lower age limit of 50 years for the age-plus risk-based scenarios given that a lower age indication is currently under review (37).

**Table 2.**
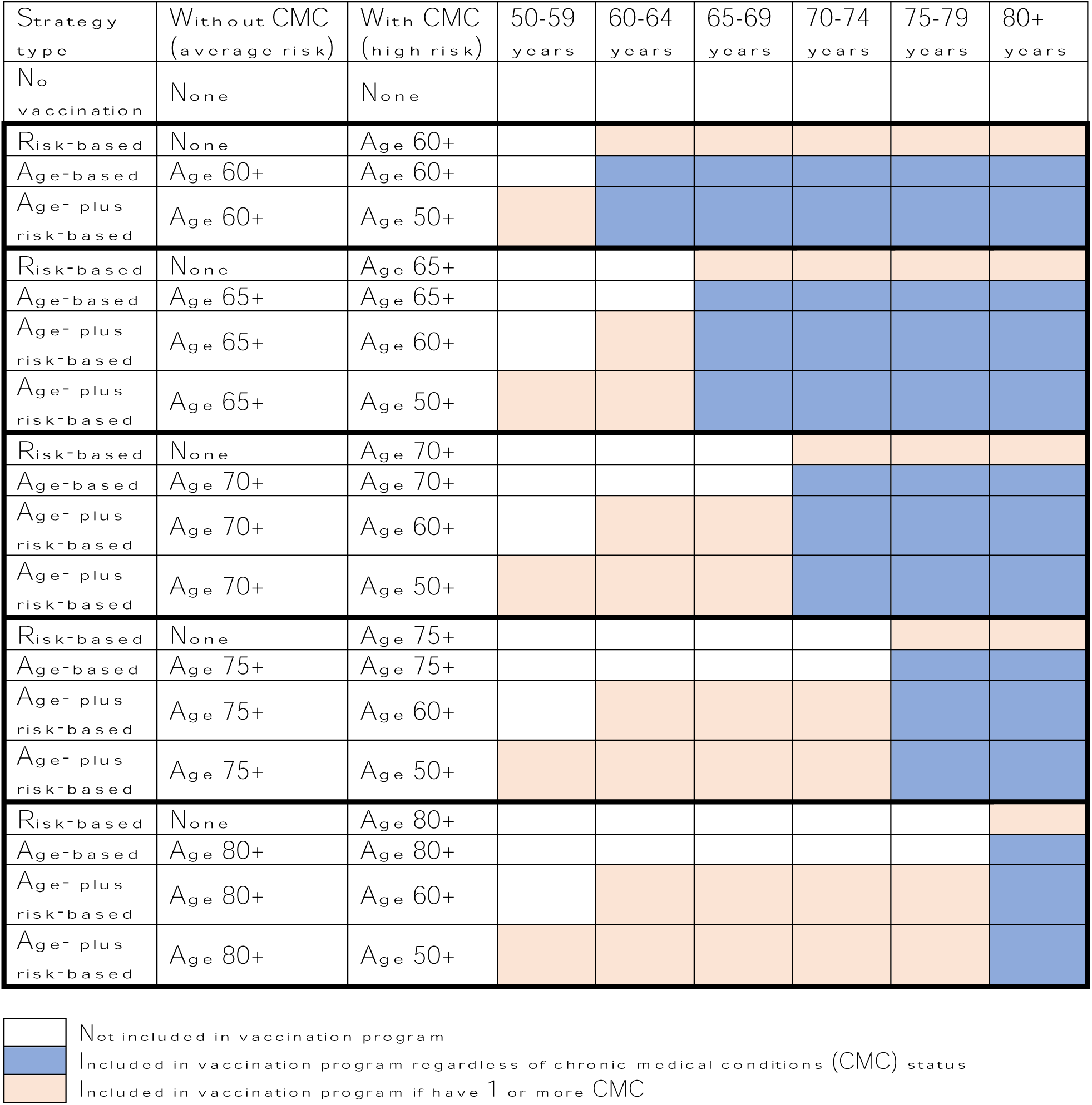
Vaccination strategies evaluated in the model.

### Model validation

We used estimates of RSV burden in adults aged 60 and older in high-income countries from a meta-analysis (3) to assess the validity of our approach for estimating RSV disease burden. Although our model includes adults aged 50 and older, we focused on the population aged 60 and older for model validation, to align with estimates from the meta-analysis. We compared our model-derived estimates of medically-attended RSV cases, hospitalizations, and deaths in the absence of vaccination to estimates for the United States population aged 60 and older (3) that were adjusted for the relative sizes of this population group in Canada and the United States.

### Analysis

Model-projected incidence of RSV treated in outpatient and inpatient settings and vaccination program costs for different vaccination strategies were used to estimate QALYs, costs, and incremental cost-effectiveness ratios (ICERs). We also calculated outcomes averted compared to no vaccination and number needed to vaccinate (NNV) to avert each modelled health outcome. Model estimates were based on 20,000 simulations (400 draws of parameters from distributions and 50 stochastic simulations per parameter set). Outcomes across strategies were compared within each model simulation and summary results across the simulations were calculated as medians and 95% credible intervals (CrI). Unless otherwise stated, results are provided for the health system perspective, with results for the societal perspective provided in the supplementary material.

### Sensitivity and scenario analyses

We performed probabilistic sensitivity analyses and generated cost-effectiveness acceptability curves to visualize the probability that competing vaccination strategies were preferred at varying cost-effectiveness thresholds.

Though the main analysis included all vaccination strategies when estimating ICERs, we also conducted sub-analyses restricted to age-based strategies only, recognizing that medical risk-based strategies may be challenging to implement.

We conducted several scenario analyses, with details provided in **Supplementary Table 1**. Briefly, we considered more optimistic scenarios for VE, including less rapid waning during season 2 or protection that extended through a third season. We also evaluated the impact of varying assumptions about the amount of under-detection of RSV disease by assuming no under-detection or more under-detection than used in the base case. We reduced the proportion of people hospitalized with RSV with one or more CMC from 98% to 90%. Finally, we evaluated the impact of RSV vaccination strategies in a setting of higher disease incidence (38) and higher costs associated with medical care, including transportation to receive medical care (39), which may reflect the context of some remote and isolated communities. We used the age distribution of the Canadian territories for this analysis, to reflect the younger age of the Northern Canadian population (15).

To address uncertainty in vaccine price, we identified the optimal strategy at different vaccine prices for different cost-effectiveness thresholds for the base case and scenario analyses. Finally, we re-estimated production losses using the friction cost approach (40) with a 3-month friction period, rather than the human capital method (41) that was used in the main analysis.

## RESULTS

### Model validation

Model-estimated cases of medically-attended RSV disease in the Canadian population aged 60 years and older for the base case analysis were consistent with expected cases derived using alternate estimates of RSV disease burden in high-income settings (**Figure 2**). Without vaccination, we projected 131,389 (95% CrI: 120,070 – 143,581) medically-attended RSV cases, 12,068 (95: CrI: 10,324 −13,883) hospitalizations, and 1,015 (95% CrI: 617-1,450) deaths annually among Canadians aged 60 years and older.

**Figure 2.**
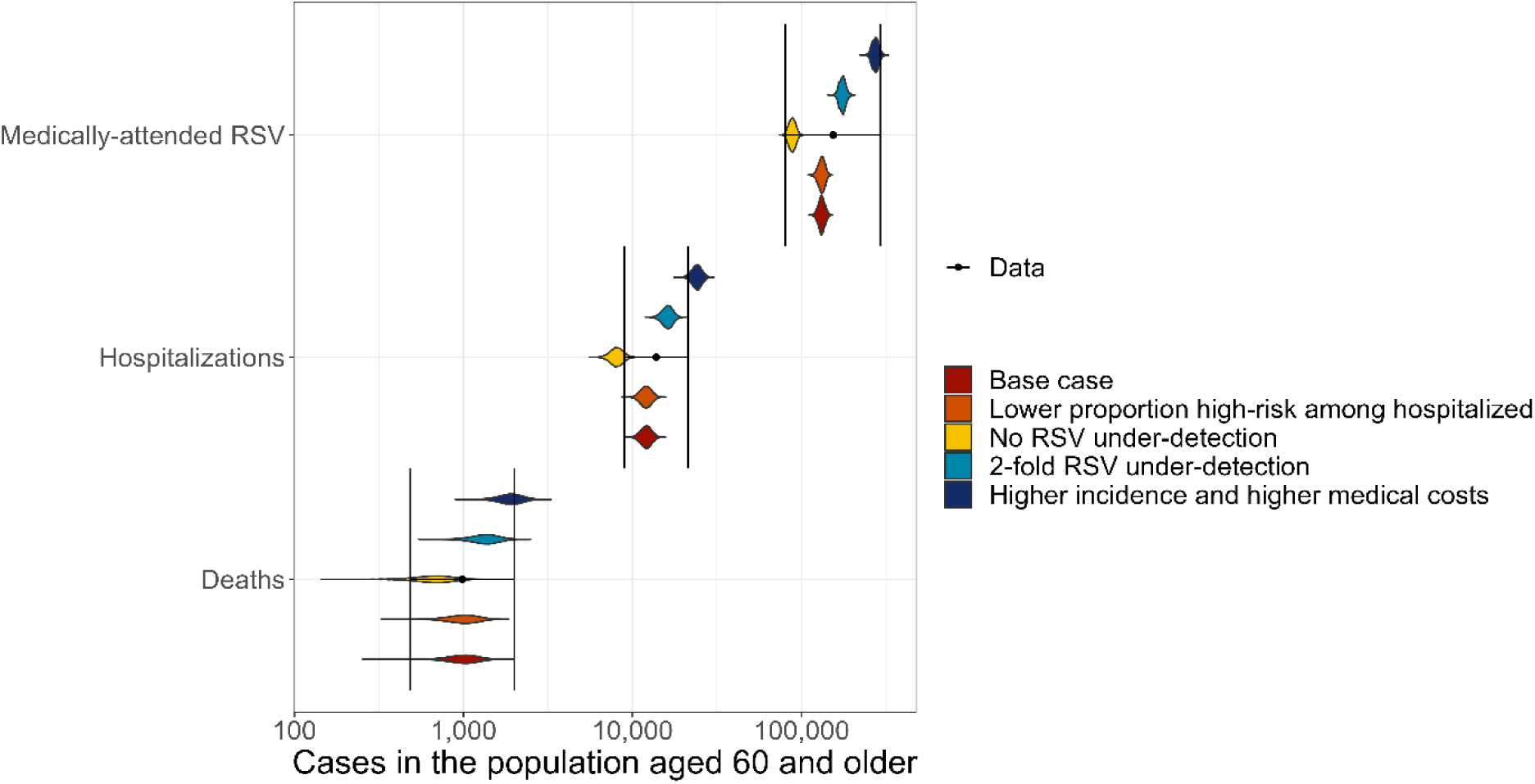
Comparison of model-projected annual RSV disease in the Canadian population aged 60 years and older to estimates from the United States, adjusted to reflect the Canadian population size. Coloured violin plots show the distribution of model estimates for the base case and scenario analyses in the absence of vaccination and black points and bars show the adjusted US estimates from Savic et al. (3). Note that results are plotted on a log scale. Scenarios that altered vaccine effectiveness assumptions are not shown, as they had no impact on the incidence of RSV outcomes in the absence of vaccination.

### Base case

For all strategies, risk reduction was largest when vaccination included younger ages (**Figure 3**, **Supplementary Table 2**). Age-based strategies were projected to avert a median of 12-30% of outpatient cases, 20-40% of hospitalized cases, and 23-41% of deaths. Using medical risk-based strategies, vaccination was projected to avert a median of 9-21% of outpatient cases, 20-39% of hospitalized cases, and 22-40% of deaths in the population, depending on the vaccine used and the assumed age recommendation. Age-plus risk-based strategies were projected to avert a median of 20-31% of outpatient cases, 38-42% of hospitalized cases, and 39-42% of deaths.

**Figure 3.**
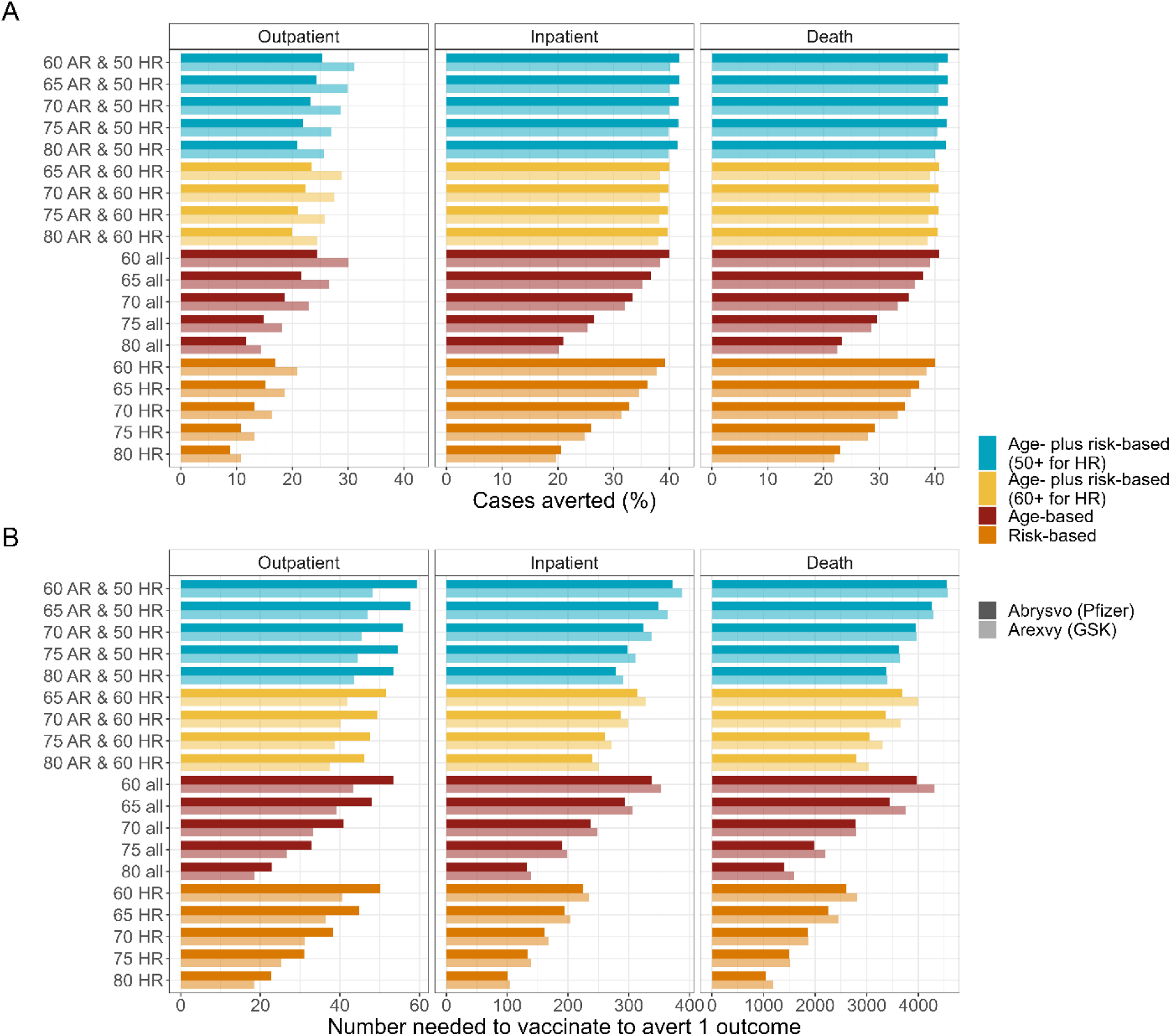
Model-projected outcomes averted and number needed to vaccinate for different RSV vaccination strategies. (A) Median values of RSV-attributable outpatient cases, inpatient cases, and deaths averted compared to no vaccination over a three-year time period. (B) Median values of number needed to vaccinate to avert one RSV-attributable outpatient case, inpatient case, or death. Note that the x-axis scales are different for the different outcomes in panel B. Colours represent the type of strategy used and shading is used to differentiate between the vaccines that were modelled. Y-axis labels indicate the age group cutoff used for the different strategies. HR = high risk (1 or more chronic medical conditions); AR = average risk (no chronic medical conditions).

Estimates of number needed to vaccinate to avert one outpatient visit, hospitalization, or death tended to be largest for the age-plus risk-based strategies and were smallest for risk-based strategies (**Figure 3, Supplementary Table 2**). For all strategies, NNV increased as the age cutoff for vaccination was lowered, though this gradient was less apparent for the age-plus risk-based strategies.

Results were not substantially different for the two vaccines evaluated. For both vaccines, a program focused on vaccinating people with at least one CMC aged 70 years and older was the optimal strategy for a cost-effectiveness threshold of $50,000 per QALY (**Figure 4**, **Table 3**). Lowering the age recommendation to people with at least one CMC aged 60 years and older resulted in ICERs of approximately $100,000 CAD per QALY gained. ICERs for age-plus risk-based strategies that used different age cutoffs depending on the presence or absence of CMCs exceeded commonly used cost-effectiveness thresholds (42, 43). No vaccination was dominated (more costly and less effective) compared to vaccination.

**Figure 4.**
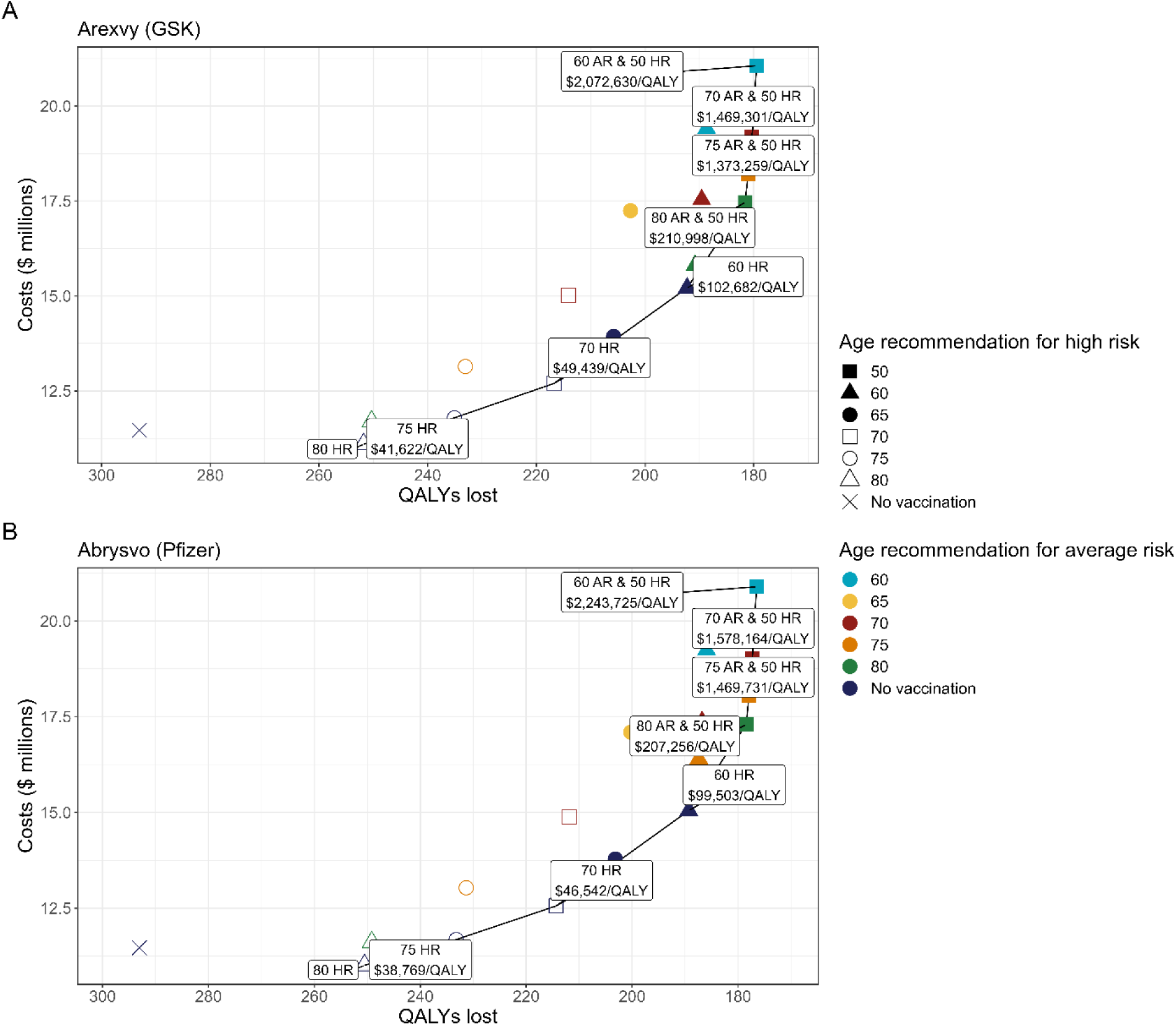
Costs and quality-adjusted life year (QALY) losses associated with each vaccination strategy. Vaccine characteristics were based on available data for (A) Arexvy (GSK) or (B) Abrysvo (Pfizer). The solid line shows the cost-effectiveness frontier, which connects strategies that are not dominated or extended dominated. Labels indicate the sequential incremental cost-effectiveness ratios (ICERs) for non-dominated strategies. Results are shown for the base case analysis for the health system perspective. Values represent the mean of 20,000 simulations per strategy. Colours indicate the age recommendation for the average-risk population and shapes indicate the age recommendation for the high-risk population. Additional details about the strategies are provided in Table 2 and costs, QALYs, and ICERs are provided in Table 3.

**Table 3.** Costs, quality-adjusted life years, and incremental cost-effectiveness ratios (ICERs) for all vaccination strategies with vaccine effectiveness estimates based on (A) Arexvy (GSK) and (B) Abrysvo (Pfizer), for the health system perspective.

**(A)**
| Strategy* | Costs (\$) | Effect (QALYs lost) | Incremental Costs (\$) | Incremental Effect (QALYs gained) | Sequential ICER (\$ per QALY) |
| --- | --- | --- | --- | --- | --- |
| 80 HR | 11,093,616 | 251.7969 | -- | -- | -- |
| 75 HR | 11,789,187 | 235.0853 | 695,572 | 16.7117 | 41,622 |
| 70 HR | 12,694,094 | 216.7820 | 904,907 | 18.3033 | 49,439 |
| 60 HR | 15,211,110 | 192.2692 | 2,517,016 | 24.5127 | 102,682 |
| 80 AR & 50 HR | 17,464,260 | 181.5907 | 2,253,150 | 10.6786 | 210,998 |
| 75 AR & 50 HR | 18,217,831 | 181.0419 | 753,571 | 0.5487 | 1,373,259 |
| 70 AR & 50 HR | 19,190,540 | 180.3799 | 972,709 | 0.6620 | 1,469,301 |
| 60 AR & 50 HR | 21,062,420 | 179.4767 | 1,871,880 | 0.9031 | 2,072,630 |
| 80 all | 11,691,842 | 250.3266 | -- | -- | Extended dominated |
| 65 HR | 13,933,048 | 205.8000 | -- | -- | Extended dominated |
| 80 AR & 60 HR | 15,809,336 | 190.7989 | -- | -- | Extended dominated |
| 75 AR & 60 HR | 16,562,907 | 190.2501 | -- | -- | Extended dominated |
| 65 AR & 50 HR | 20,176,398 | 179.9373 | -- | -- | Extended dominated |
| No vaccination | 11,462,844 | 293.0558 | -- | -- | Dominated |
| 75 all | 13,140,984 | 233.0662 | -- | -- | Dominated |
| 70 all | 15,018,600 | 214.1009 | -- | -- | Dominated |
| 65 all | 17,243,412 | 202.6763 | -- | -- | Dominated |
| 70 AR & 60 HR | 17,535,616 | 189.5881 | -- | -- | Dominated |
| 65 AR & 60 HR | 18,521,474 | 189.1455 | -- | -- | Dominated |
| 60 all | 19,407,496 | 188.6850 | -- | -- | Dominated |

**(B)**
| Strategy* | Costs (\$) | Effect (QALYs lost) | Incremental Costs (\$) | Incremental Effect (QALYs gained) | Sequential ICER (\$ per QALY) |
| --- | --- | --- | --- | --- | --- |
| 80 HR | 11,005,219 | 250.5372 | -- | -- |  |
| 75 HR | 11,677,018 | 233.2090 | 671,800 | 17.3283 | 38,769 |
| 70 HR | 12,555,211 | 214.3400 | 878,192 | 18.8690 | 46,542 |
| 60 HR | 15,048,253 | 189.2850 | 2,493,042 | 25.0550 | 99,503 |
| 80 AR & 50 HR | 17,295,502 | 178.4421 | 2,247,249 | 10.8429 | 207,256 |
| 75 AR & 50 HR | 18,049,620 | 177.9290 | 754,117 | 0.5131 | 1,469,731 |
| 70 AR & 50 HR | 19,022,818 | 177.3123 | 973,198 | 0.6167 | 1,578,164 |
| 60 AR & 50 HR | 20,895,542 | 176.4777 | 1,872,724 | 0.8346 | 2,243,725 |
| 80 all | 11,603,970 | 249.1642 | -- | -- | Extended dominated |
| 65 HR | 13,781,691 | 203.1625 | -- | -- | Extended dominated |
| 80 AR & 60 HR | 15,647,004 | 187.9120 | -- | -- | Extended dominated |
| 75 AR & 60 HR | 16,401,122 | 187.3989 | -- | -- | Extended dominated |
| 65 AR & 50 HR | 20,009,151 | 176.9007 | -- | -- | Extended dominated |
| No vaccination | 11,462,844 | 293.0558 | -- | -- | Dominated |
| 75 all | 13,029,887 | 231.3228 | -- | -- | Dominated |
| 70 all | 14,881,277 | 211.8372 | -- | -- | Dominated |
| 65 all | 17,094,090 | 200.2481 | -- | -- | Dominated |
| 70 AR & 60 HR | 17,374,320 | 186.7822 | -- | -- | Dominated |
| 65 AR & 60 HR | 18,360,653 | 186.3706 | -- | -- | Dominated |
| 60 all | 19,247,044 | 185.9475 | -- | -- | Dominated |

\*HR = high risk (one or more chronic medical conditions); AR = average risk (no chronic medical conditions)

Results for the societal perspective were qualitatively similar regardless of whether the human capital (**Supplementary Table 3**) or friction cost (**Supplementary Table 4**) approach was used for estimating production losses, though the estimated ICERs were lower than those for the health system perspective.

Probabilistic sensitivity analysis identified some uncertainty about the optimal strategy around the $50,000 per QALY threshold, with the risk-based strategy most likely to be cost-effective shifting from age 80 to age 70 for both vaccines near this threshold (**Supplementary Figure 2**). Risk-based vaccination of people aged 70 and older had the largest probability of being cost-effective from $50,000 up to to a threshold of $80,000 per QALY. Beyond $80,000 per QALY, there was a less clear difference between the age 60 and age 70 risk-based strategies.

Age-only strategies were never identified as cost-effective options regardless of the cost-effectiveness threshold used, when considered alongside other strategies. When only evaluating age-based strategies, vaccinating adults aged 80 years and older resulted in sequential ICERs of $3,261-5,391 per QALY gained. Lowering the age recommendation for all adults to 75 years and older required a cost-effectiveness threshold of approximately $80,000 per QALY (**Supplementary Table 5**).

For the base case and a cost-effectiveness threshold of $50,000 per QALY, a 40% reduction in vaccine price per dose (to $135-140 from the list price of $230) was needed for the age cutoff for the optimal strategy to change from risk-based aged 70 and older, to risk-based aged 60 and older (**Figure 5**).

**Figure 5.**
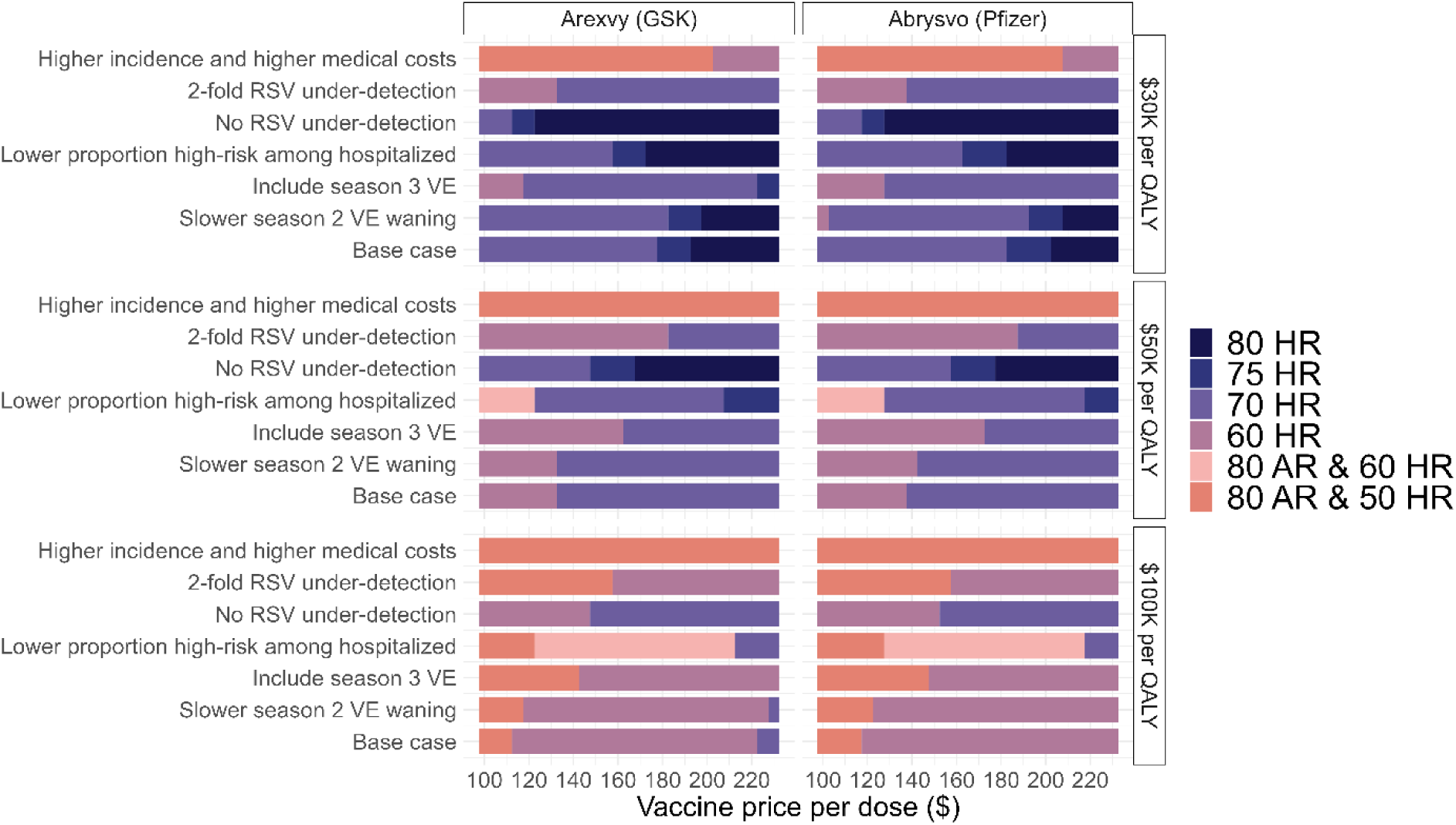
Impact of vaccine price on optimal vaccination strategy for different scenarios and cost-effectiveness thresholds. For a given vaccine price per dose (x-axis) the optimal vaccination strategy is shown for cost-effectiveness thresholds of $30,000, $50,000, or $100,000 per QALY. The base case vaccine price was $230 per dose (maximum value on the x-axis). Scenario details are provided in Supplementary Table 1. Results are shown for the health system perspective for the indicated vaccines. HR = high risk (one or more chronic medical conditions); AR = average risk (no chronic medical conditions).

### Scenario analyses

While most of the scenario analyses generated estimates of RSV outcomes that were compatible with expected RSV burden, the no RSV under-detection scenario appeared to under-estimate burden, while the higher incidence and higher medical costs scenario over-estimated burden (**Figure 2**).

Medical risk-based strategies were generally preferred across most scenarios, except with substational vaccine price reductions and a higher cost-effectiveness threshold (**Figure 5**). However, in the higher incidence and higher medical costs scenario, age-plus medical-risk stategies were optimal even at higher vaccine prices and lower thresholds. At vaccine list prices and a $50,000 per QALY threshold, an older age cutoff for risk-based strategies was optimal in scenarios assuming a lower proportion of people with CMCs among hospitalized cases or no underdetction of RSV disease.

Assumptions that vaccine protection either extended into a third season or waned more slowly in the second season had little impact on results. For both scenarios, at the vaccine list prices, vaccinating high-risk adults aged 70 years and older remained optimal at a $50,000 per QALY threshold. If vaccine protection extends through a third RSV season, a 25% reduction in vaccine price (to $165-175 per dose) would be required for the age recommendation for a risk-based strategy to be lowered to from 70 to 60 years and older.

In a sub-analysis of age-based strategies only, we observed similar trends as for the full analysis, with a lower age cutoff preferrable when the cost-effectiveness threshold was increased and/or the vaccine price was lowered (**Supplementary Figure 3**).

## INTERPRETATION

Our model-based cost-utility analysis of RSV vaccination for the Canadian population shows that strategies focused on adults with underlying medical conditions that place them at increased risk of RSV disease are more likely to be cost-effective than general age-based strategies. We found that vaccination of older adults may be less costly and more effective than no vaccination and that vaccinating people aged 70 years and older with CMCs is likely to be cost-effective based on commonly used cost-effectiveness thresholds. Our finding that medical risk-based policies were preferred over age-based ones were robust to a range of alternate assumptions, including vaccine protection that extends into a third RSV season or a lower proportion of people with CMCs among hospitalized cases. Our results were sensitive to assumptions about vaccine price, but risk-based approaches were preferred even at lower vaccine prices. We found that broader programs may be cost-effective in settings where the risk of disease is higher and healthcare costs are higher, such as some remote communities in Northern Canada.

Age-based strategies were never cost-effective compared to risk-based or age plus risk-based strategies, regardless of the cost-effectiveness threshold used or the vaccine price considered; age-based strategies were either dominated (i.e., more costly and less effective) or extendedly dominated (i.e., there was a combination of other strategies that would result in more QALYs gained at lower costs). While age-based strategies would not result in the optimal use of resources when risk-based vaccination strategies are an option, we did a sub-analysis evaluating only age-based strategies because there may be other reasons why such an approach would be desirable, such as ease of identification of people recommended for vaccination by healthcare providers. Of note, our vaccination coverage estimates, which were based on influenza vaccination, assumed that for all age groups, uptake is higher for people with CMCs; as such, even the age-based strategies we evaluated include an element of risk-focused vaccination.

A recent review identified five economic evaluations for RSV vaccines in high-income countries (excluding Canada) (11), all of which evaluated age-based strategies only in the population aged 60 or 65 years and older. Without a signficant reduction in vaccine price, all of the non-industry funded analyses estimated ICERs exceeding $100,000 per QALY gained (44–46). Similarly, a Canadian economic evaluation (47) assessed the vaccine price required for an RSV vaccination program to be cost-effective at a threshold of $50,000 per QALY and found that substantial vaccine price reductions would be required to be cost-effective for use in the general population. By contrast, in this same study, smaller price reductions were required for the vaccine to be cost-effective for a program for residents of long-term care homes (47). Overall, these findings are consistent with our analysis, suggesting a risk-focused vaccination program may be optimal.

Our model-based analysis has several limitations. In the absence of data showing that RSV vaccines prevent onward transmission following infection, we used a static model to estimate the impact of RSV vaccination programs. Absent indirect effects, our estimates of cost-effectiveness of RSV vaccination programs may be overly conservative, though a recent dynamic model showed that assumptions about VE for reducing transmission are not expected to substantially influence the estimated impact of RSV vaccination programs in older adults (48). Vaccine effectiveness and waning assumptions were based on data for two RSV seasons, and we assumed a single VE for all ages and risk groups, given available data. As additional data on durability of vaccine protection and VE in different population groups accumulate, we can refine our estimates of vaccination program impact. In particular, if VE is found to be reduced with increasing age or presence of CMCs, the preference for risk-based strategies may be diminished. The finding that risk-based strategies are optimal is informed by available data showing higher incidence of severe RSV disease in people with underlying medical conditions. In our analysis, estimates of the proportion of the population with one or more CMCs were based on underlying health conditions that could place individuals at elevated risk of complications following SARS-CoV-2 infection (16), which may not align with risk of medically-attended RSV disease. In general, additional data on the burden of RSV disease in Canadian adults, particularly in the outpatient setting, would contribute to an improved understanding of the potential benefits of vaccination programs.

In summary, based on currently available data, RSV vaccination programs in some groups of older Canadians are expected to be cost-effective, with programs focusing on people with underlying medical conditions that place them at increased risk of severe RSV disease expected to provide the best value for money.

## Data Availability

All data produced in the present work are contained in the manuscript

## Acknowledgements

The authors thank Ruoke Chen from the Public Health Agency of Canada for providing estimates of influenza vaccination coverage and members of the National Advisory Committee on Immunization RSV Working Group for providing feedback during model development.

