## Supplementary Material, Figures, and Tables for "Respiratory syncytial virus vaccination strategies for older Canadian adults: a cost-utility analysis"

**Model technical appendix**

*Model overview*

We built a static individual-based model of medically-attended RSV disease to explore the impact of alternate age-, medical risk- and age- plus medical risk-based vaccination policies on RSV-associated outcomes. The model followed a multi-age population cohort of 100,000 people over a three-year period that included three full RSV seasons, with the age group distribution based on projections of the Canadian population aged 50 and older (1). It was a closed population cohort but individuals could die, either due to RSV-attributable mortality or mortality from other causes. Given the three-year time horizon of the model, individuals could also age into the next oldest age grouping between each RSV season, with a probability of 1/size of the current age grouping; individuals in the oldest age grouping remained there. Individuals were further characterized by the presence or absence of one or more chronic medical conditions (CMCs) (2). The model was assumed to start in September, at the start of a new RSV season, and used monthly time steps. A portion of the population was vaccinated in the first two months of model entry with vaccination coverage based on influenza vaccine uptake (3). We included risks of solicited severe local and severe systemic adverse events following immunization. Unsolicited serious adverse events were not included. RSV infection could occur at any month during the three-year period and was assumed to follow seasonal trends, with peak activity occurring from January to March. We modelled medically-attended RSV disease only, with individuals with RSV disease requiring one of the following levels of care: healthcare provider visit, emergency department visit, or hospitalization, with or without ICU admission (**Error! Reference source not found.**). RSV-attributable death occurred only among people hospitalized with RSV. Vaccination was assumed to reduce the risk of these outcomes. Although multiple RSV infections are possible within a season and across seasons (4), for simplicity, we assumed a maximum of one of medically-attended RSV disease event per person over the model time horizon.

The model was static and did not incorporate dynamic feedbacks, such as indirect effects of vaccination in unvaccinated individuals. A three-year period was used based on currently available data showing that vaccine protection lasts for at least two RSV seasons. The three-year period allowed for the investigation of vaccine protection that lasts through three RSV seasons in a scenario analysis, though the base case analysis assumed vaccine protection for two seasons only. A lifetime time horizon was used at the individual level to capture the costs and consequences of RSV-attributable mortality. Costs and QALYs were used to calculate ICERs. We also calculated number needed to vaccinate (NNV) to avert each modelled health outcome. Costs are in 2023 Canadian dollars and, where necessary, were converted using the Canadian Consumer Price Index (5). A discount rate of 1.5% was used for costs and outcomes and cost-effectiveness was assessed from both the health system and societal perspectives (6). The model was constructed and analyzed using R (7). The study followed the ISPOR Consolidated Health Economic Evaluation Reporting Standards (CHEERS) 2022 reporting guidance.

Model parameters describing RSV epidemiology, vaccine characteristics, costs, and health utilities (**Table 1**) were obtained from published studies and available data, when possible, and by assumption or expert opinion otherwise. Canadian data were used preferentially and we used age- and CMC-status specific estimates, when available. In the tables, parameters with ranges included indicate parameters that were drawn from distributions for the analysis, with beta distributions used for probabilities and proportions and utilities and gamma distributions used for costs.

*RSV epidemiology*

RSV seasonality was modelled by assuming that annual incident infections were distributed over the months of the year. The proportion of annual cases occurring each month was estimated from reported RSV tests and positive detections for all of Canada reported by the Respiratory Virus Detection Surveillance System for nine seasons (2010-2011 to 2018-2019) (8). We assumed that RSV seasonality followed patterns observed prior to the COVID-19 pandemic.

Seasonal incidence of hospitalized RSV and risk of ICU admission were obtained from a Canadian study (9). Seasonal RSV hospitalization incidence was adjusted to annual incidence using the reported proportion of RSV cases reported between November and May (the time period used to estimate seasonal hospitalization rates) (8).

In the absence of Canadian estimates of outpatient incidence of medically-attended RSV disease, we used data from a meta-analysis that estimated rates of hospitalization, ED admissions, and outpatient healthcare provider visits (excluding ED admissions) in the United States (10) to estimate incidence of these outcomes. We applied the ratio of rates of ED or healthcare provider visits compared to hospitalized cases from the meta-analysis to the Canadian hospitalization rates. RSV-attributable mortality was restricted to hospitalized cases. RSV case-fatality rates were adjusted for background mortality due to other causes. We accounted for the association of comorbidities with increased risk of medically-attended RSV disease (11, 12) by adjusting age-specific outpatient and inpatient incidence estimates to be consistent with the observed fractions of people receiving outpatient care or hospitalized with RSV with at least one chronic medical condition (9, 11).

A recent systematic review and meta-analysis identified that RSV disease in adults is likely under-detected, with detection approximately 1.5-times higher than is detected by commonly used approaches (10). We adjusted all RSV outcome estimates by this under-detection ratio in the base case analysis. We evaluated no under-detection and a higher under-detection factor in scenario analyses.

*Vaccine characteristics*

We used data for the two RSV vaccines (Arexvy (GSK), and Abrysvo (Pfizer)) currently authorized for use in Canada to estimate vaccine effectiveness (VE) and rates of adverse events following immunization (AEFIs) (13-16). As the endpoints measured varied across vaccine products, we used the following general classification to assign VE estimates from randomized-controlled trials to the modelled health outcomes: for outpatient care, we used VE for medically-attended RSV-related lower respiratory tract disease (LRTD) (Arexvy) or VE for medically-attended lower respiratory tract illness (LRTI)-RSV with two or more signs or symptoms (Abrysvo); for hospitalization, we used VE for severe RSV-related LRTD (Arexvy) or VE for medically-attended LRTI-RSV with 3 or more signs or symptoms (Abrysvo).

Vaccine protection began the month following vaccination, such that individuals vaccinated in September had maximum protection in October, and those vaccinated in October had maximum protection in November. Data on average VE and average duration of follow-up for each season were used to generate step functions, with protection assumed to wane linearly between seasons. In the absence of data for season 3, we assumed that VE reached one-third of season 2 VE by the end of the season (i.e., month 36). We used a cubic polynomial regression model to obtain smoothed estimates of VE over 36 months (**Supplementary Figure 1**). We assumed that VE did not vary by age or CMC status. Since RCT data were only available for up to two RSV seasons, in our base case analysis we conservatively assumed that VE in season 3 was 0. We modelled VE extending through to season 3 in a scenario analysis.

*Costs*

Costs of inpatient RSV were based on attributable costs derived from a retrospective population-based cohort study in Ontario, Canada (17). Costs for outpatient cases were based on estimates for influenza (18). Vaccination costs included administration costs and public Canadian list prices. Direct costs for AEFIs included a healthcare provider visit and treatment costs (18-20).

For the societal perspective, costs included patient productivity loss due to AEFIs and RSV-attributable illness and death, caregiver productivity loss, and out-of-pocket medical costs. Productivity loss was estimated using the human capital method (21) in the main analysis. Age-specific labour force participation rates (22) and average employment income were obtained from Statistics Canada (23). Caregiver wages were estimated based on the average employment income and labour force participation of the population aged 25 to 54 years (23). The amount of caregiver productivity loss was assumed to be equivalent to length of illness multiplied by average reduction in productivity for caregivers (24).

*Utilities*

Age-specific utilities for the general population were based on EQ-5D-5L index scores for the Canadian population (25). QALY losses associated with the modelled health outcomes were derived from published studies and assumption (26-30).

*Vaccination strategies*

We evaluated a combination of age-only, medical risk-only, and age plus medical risk-based strategies (**Table 2**). All strategies assumed administration of one dose of vaccine. For age-based strategies, all individuals aged greater than or equal to the specified age cutoff (i.e., 60, 65, 70, 75, or 80 years and older) were eligible to receive the vaccine. For medical risk-based strategies, only individuals aged greater than or equal to the specified age cutoff who also had one or more CMCs were eligible to receive the vaccine. For age plus medical risk-based strategies, there were two age thresholds: one applied universally to all individuals above the specified age cutoff; a second, lower age cutoff applied to individuals with at least one CMC who were younger than the universal age requirement. We evaluated a lower bound for people with CMC of either 50 years or 60 years for the age- plus medical risk-based-strategies. Although the vaccines are currently authorized for use in adults aged 60 and older, we considered a lower age limit of 50 years for the age-plus medical risk-based scenarios given that a lower age indication is currently under review (31). Recognizing that medical risk-based strategies may be challenging to implement, we conducted a sub-analysis that was restricted to age-based strategies only.

*Model validation*

We used estimates of RSV burden in adults aged 60 and older in high-income countries from a meta-analysis (32) to assess the validity of our approach to estimating RSV disease burden. Although our model includes adults aged 50 and older, we focused on the population aged 60 and older for model validation to align with the population estimates available in the systematic review, which only included adults aged 60 and older. Specifically, we used estimates of RSV-associated acute respiratory infections (1.21 million, 95% CI: 0.63-2.31 million), hospitalizations (108,834, 95% CI: 70,555-168,130), and in-hospital deaths (7,763, 95% CI: 3,813-15,739) for the United States population aged 60 and older (32) and adjusted these estimates for the relative sizes of this population group in Canada (1) and the United States (33) (with the US population approximately 8-times the size of the Canadian population for this age group). Model-based estimates of medically-attended (MA)-RSV cases, hospitalizations, and deaths per 100,000 population were converted to absolute numbers for the population aged 60 and older. These two values were compared to determine if the model estimates were consistent with estimates from another high-income country. MA-RSV estimates from our model were calculated as the sum of all RSV outpatient and inpatient cases.

*Analysis*

In the individual-based model, modelled individuals are assigned characteristics, such as age group, CMC status, and vaccination status. Outcomes are generated for each individual and summarized across the modelled population. These types of models incorporate random chance (i.e., are stochastic, first-order uncertainty), such that each time the model is simulated, we will obtain a different distribution of outcomes. To explore uncertainty in the model parameters (second-order uncertainty), parameters were drawn from distributions, and each parameter set was used to simulate the vaccination scenarios for 50 model simulations (to capture first-order uncertainty). Probabilistic model estimates were based on 20,000 simulations (400 second-order and 50 first-order). Outcomes across strategies were compared within each model simulation and summary results across the simulations were calculated as medians and 95% credible intervals (CrI).

A sequential analysis was conducted to compare ICERs for the vaccination strategies. In the sequential analysis, a strategy is eliminated if there are other strategies that are projected to result in more QALYs gained at lower costs (i.e., the strategy is dominated) or there is a combination of other strategies that would result in more QALYs gained at lower costs, such that the excluded strategy would never be the optimal intervention, regardless of the cost-effectiveness threshold used (i.e., the strategy is subject to extended dominance).

*Sensitivity and scenario analyses*

Cost-effectiveness acceptability curves were generated from the probabilistic outputs to assess the probability of the vaccination strategies being cost-effective at various cost-effectiveness thresholds.

We conducted several scenario analyses, with details provided in **Supplementary Table 1**. Briefly, we considered more optimistic scenarios for vaccine production, including less rapid waning during season 2 and protection that extended through a third season. We also evaluated the impact of assumptions about the amount of under-detection of RSV cases by assuming no under-detection or a higher value than used in the base case analysis. The estimated proportion of people hospitalized with RSV who have one or more CMC was 98% in the base case analysis (9). In a scenario analysis, we reduced this proportion to 90%. Finally, a scenario analysis was conducted to evaluate the impact of RSV vaccination strategies in a setting of higher disease incidence and higher costs associated with medical care, including transportation to receive medical care, which may reflect the context of some remote and isolated communities. Burden of RSV in older adults is less well characterized than in children; we based our estimates of increased burden on hospitalization rates for respiratory infections (34). Relative rates were applied to the base case estimates of inpatient and outpatient RSV incidence. Relative increases in costs for inpatient and outpatient medical care were based on data for pneumococcal disease and applied to base case costs (35). We used the age distribution of the Canadian territories for this analysis, to reflect the younger age of the Northern population (1).

To address uncertainty in vaccine price, we identified the optimal strategy at different vaccine prices for different cost-effectiveness thresholds for the base case and all scenario analyses. We re-estimated production losses using the friction cost approach. In contrast with the human capital approach, the friction cost approach assumes that after a “friction period”, workers who have left the workforce will eventually be replaced by currently unemployed workers or existing employed workers (36). We used a three-month friction period for people who died of RSV (37).

### Supplementary Figures

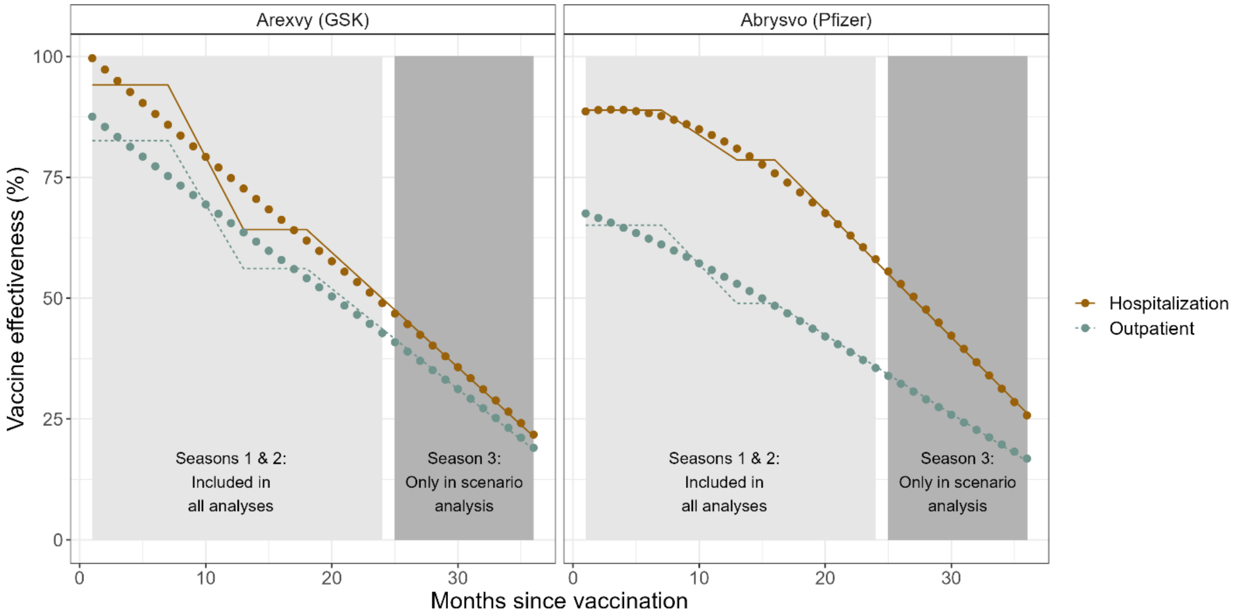


**Supplementary Figure 1.** Estimated monthly vaccine effectiveness (VE). Lines show the VE modelled as step functions, based on average VE and follow-up times for the two RSV seasons. Points show the smoothed estimates used in the model. VE for season 3 is based on assumption. In the base case analysis, VE was assumed to be zero for season 3; VE extending into season 3 was considered in a scenario analysis.

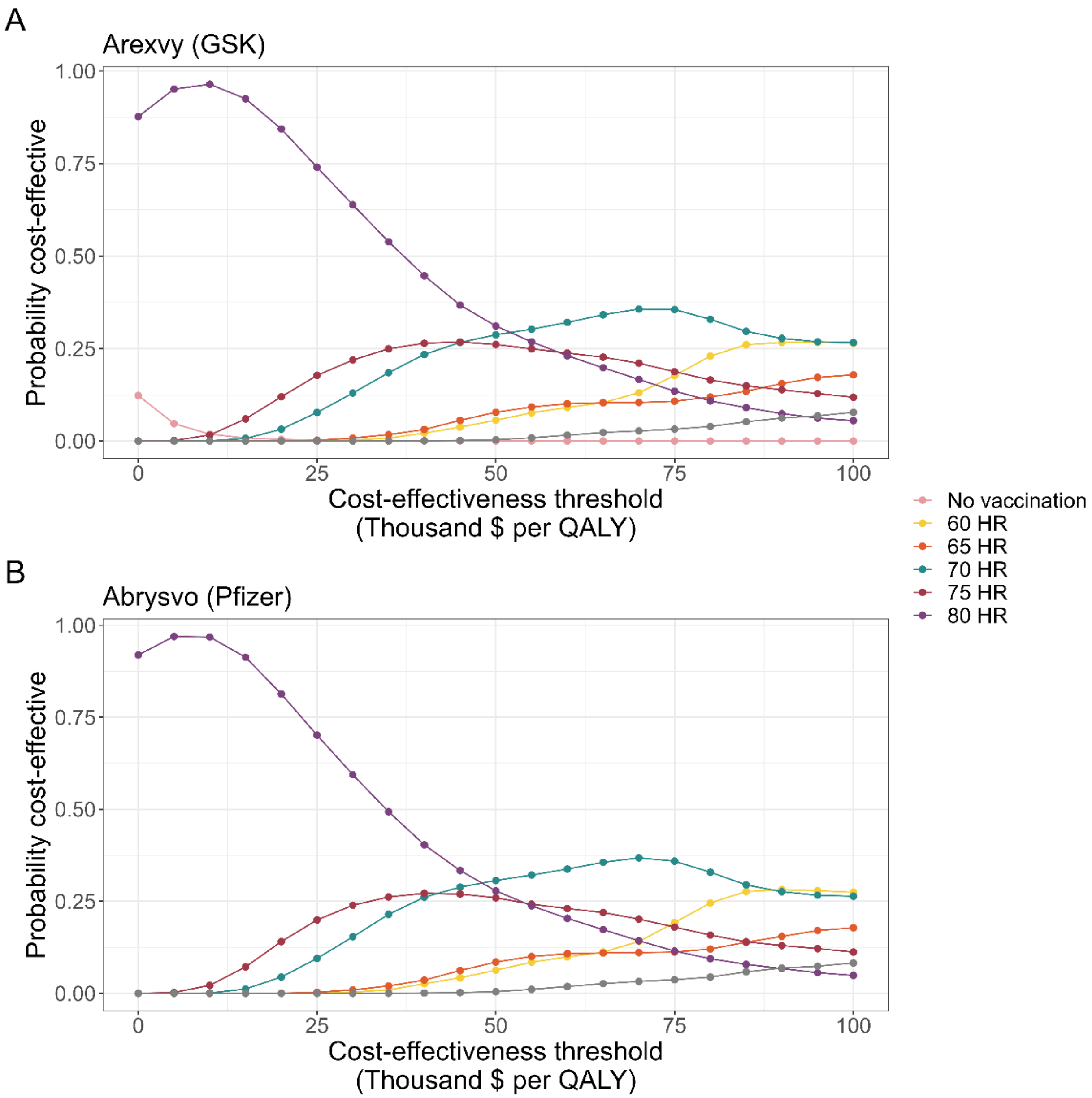


**Supplementary Figure 2**. Proportion of simulations for which each vaccination strategy was the optimal strategy for a given cost-effectiveness threshold. Results are shown for vaccine characteristics based on (A) Arexvy (GSK) and (B) Abrysvo (Pfizer) and are based on 20,000 model simulations per strategy from the health system perspective. Strategies with probabilities less than 0.1 for all cost-effectiveness thresholds are not displayed to help with interpretability.

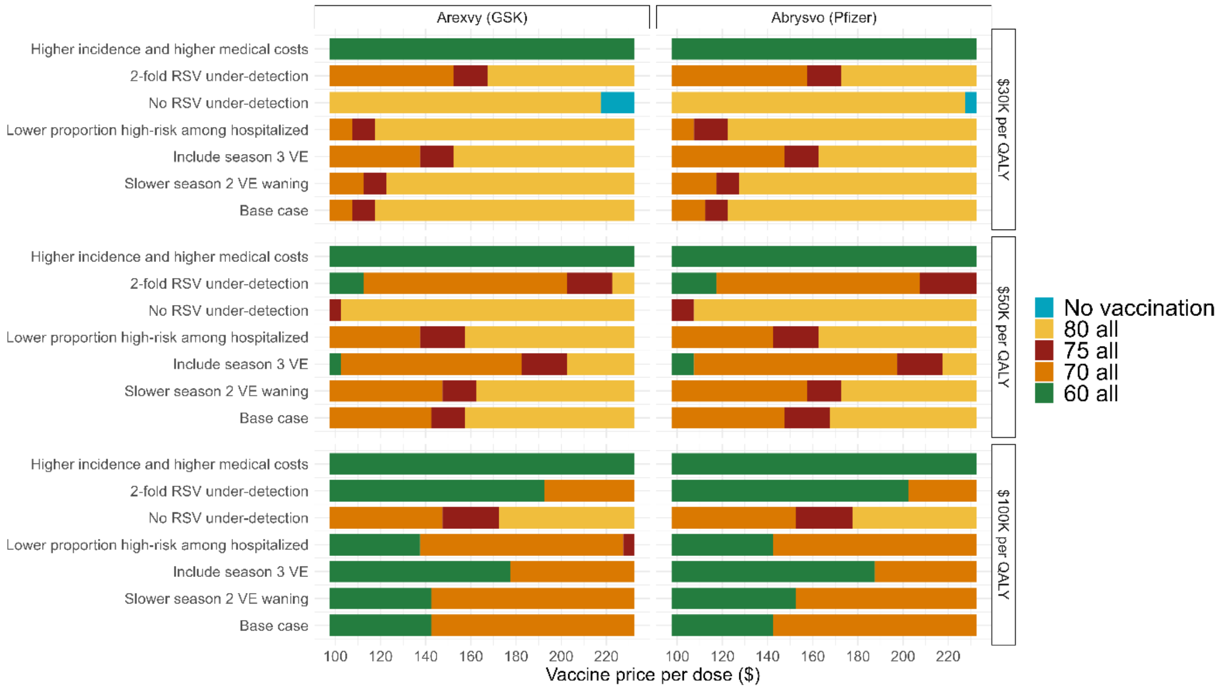


**Supplementary Figure 3**. Impact of vaccine price on optimal age-based vaccination strategy for different scenarios and cost-effectiveness thresholds. For a given vaccine price per dose (x-axis) the optimal vaccination strategy is shown for cost-effectiveness thresholds of $30,000, $50,000, or $100,000 per QALY. The base case vaccine price was $230 per dose (maximum value on the x-axis). Scenario details are provided in Supplementary Table 1. Results are shown for the health system perspective for the indicated vaccines. Only age-based strategies were included in this sub-analysis.

### Supplementary Tables

**Supplementary Table 1.** Scenario analyses.

| Scenario description | Base case | Scenario |
|---|---|---|
| Less rapid waning of VE in season 2 | VE estimates for season 2 are the average values for 4-7 months of follow up, as reported in the RCTs | VE estimates for season 2 are the average value for 12 months |
| VE extends through season 3 | VE is 0 in season 3 | VE wanes to one-third of the average season 2 VE estimate by the end of season 3 |
| No or more under-detection of outpatient and inpatient RSV cases | RSV burden is 1.5-times higher than reported estimates | There is: (i) no under-detection of RSV burden; or (ii) RSV burden is 2-times higher than reported estimates |
| Lower proportion of people with CMC among inpatient RSV cases | 98% of people hospitalized with RSV have one or more CMC | 90% of people hospitalized with RSV have one or more CMC |
| Higher RSV burden and higher medical costs | Population age distribution: based on entire Canadian population | Outpatient medical care costs (including travel): 2-times base value (35)<br>Inpatient medical care costs: 1.2-times base case value (35)<br>Inpatient medical care transport: 19-time base case value (35)<br>RSV incidence: 3-times base case values (34)<br>Population age distribution: based on population in Canadian Territories (1) |

**Supplementary Table 2.** Model-projected cases, risk reduction, and number needed to vaccinate (median and 95% credible intervals).

| | | Cases per 100,000 person-years | | | Risk reduction (%) | | | Number needed to vaccinate | | |
|---|---|---|---|---|---|---|---|---|---|---|
| **Strategy*** | **Vaccine** | **Outpatient** | **Inpatient** | **Death** | **Outpatient** | **Inpatient** | **Death** | **Outpatient** | **Inpatient** | **Death** |
| No vaccination | No vaccination | 1252<br>(1141 - 1373) | 121<br>(104 - 138) | 10<br>(7 - 14) | 0<br>(0 - 0) | 0<br>(0 - 0) | 0<br>(0 - 0) | -- | -- | -- |
| 60 HR | GSK | 990<br>(901 - 1088) | 75<br>(63 - 88) | 6<br>(3 - 9) | 21<br>(19 - 22) | 38<br>(33 - 43) | 38<br>(20 - 57) | 41<br>(36 - 46) | 234<br>(193 - 290) | 2816<br>(1641 - 6200) |
| | Pfizer | 1039<br>(946 - 1142) | 73<br>(62 - 86) | 6<br>(3 - 9) | 17<br>(16 - 18) | 39<br>(34 - 44) | 40<br>(22 - 59) | 50<br>(45 - 57) | 224<br>(186 - 278) | 2603<br>(1626 - 6176) |
| 65 HR | GSK | 1019<br>(927 - 1120) | 79<br>(67 - 92) | 7<br>(3 - 10) | 19<br>(17 - 20) | 35<br>(29 - 40) | 36<br>(18 - 54) | 36<br>(32 - 41) | 204<br>(167 - 255) | 2460<br>(1380 - 6105) |
| | Pfizer | 1063<br>(967 - 1167) | 77<br>(65 - 90) | 6<br>(3 - 10) | 15<br>(14 - 16) | 36<br>(31 - 41) | 37<br>(20 - 56) | 45<br>(40 - 51) | 195<br>(161 - 243) | 2253<br>(1369 - 4962) |
| 70 HR | GSK | 1048<br>(953 - 1153) | 83<br>(70 - 96) | 7<br>(4 - 10) | 16<br>(15 - 18) | 31<br>(26 - 37) | 33<br>(16 - 52) | 31<br>(27 - 36) | 168<br>(136 - 214) | 1875<br>(1098 - 4636) |
| | Pfizer | 1086<br>(989 - 1194) | 81<br>(69 - 94) | 7<br>(4 - 10) | 13<br>(12 - 15) | 33<br>(27 - 38) | 35<br>(17 - 53) | 38<br>(34 - 44) | 161<br>(131 - 205) | 1853<br>(1086 - 4618) |
| 75 HR | GSK | 1086<br>(988 - 1195) | 91<br>(77 - 105) | 7<br>(4 - 11) | 13<br>(12 - 15) | 25<br>(20 - 30) | 28<br>(12 - 46) | 25<br>(22 - 29) | 140<br>(110 - 182) | 1519<br>(816 - 4062) |
| | Pfizer | 1117<br>(1017 - 1228) | 89<br>(76 - 103) | 7<br>(4 - 11) | 11<br>(9 - 12) | 26<br>(21 - 31) | 29<br>(13 - 47) | 31<br>(27 - 36) | 133<br>(106 - 174) | 1502<br>(810 - 4039) |
| 80 HR | GSK | 1116<br>(1015 - 1228) | 97<br>(83 - 112) | 8<br>(4 - 11) | 11<br>(9 - 12) | 20<br>(15 - 25) | 22<br>(8 - 39) | 18<br>(16 - 22) | 105<br>(80 - 142) | 1194<br>(597 - 3638) |
| | Pfizer | 1142 | 96 | 8 | 9 | 21 | 23 | 23 | 100 | 1048 |

| | | | | | | | | | | |
|---|---|---|---|---|---|---|---|---|---|---|
| | | (1040 - 1255) | (82 - 111) | (4 - 11) | (8 - 10) | (16 - 26) | (8 - 40) | (19 - 27) | (77 - 136) | (563 - 3622) |
| 60 all | GSK | 876<br>(797 - 964) | 75<br>(63 - 87) | 6<br>(3 - 9) | 30<br>(28 - 32) | 38<br>(33 - 44) | 39<br>(21 - 58) | 43<br>(39 - 49) | 353<br>(292 - 436) | 4315<br>(2505 - 9506) |
| | Pfizer | 946<br>(861 - 1040) | 73<br>(61 - 85) | 6<br>(3 - 9) | 24<br>(23 - 26) | 40<br>(35 - 45) | 41<br>(23 - 59) | 53<br>(48 - 60) | 338<br>(281 - 418) | 3973<br>(2489 - 9456) |
| 65 all | GSK | 919<br>(835 - 1012) | 78<br>(66 - 91) | 6<br>(3 - 10) | 27<br>(25 - 28) | 35<br>(30 - 40) | 36<br>(19 - 55) | 39<br>(35 - 44) | 306<br>(251 - 382) | 3756<br>(2103 - 7606) |
| | Pfizer | 982<br>(893 - 1080) | 76<br>(65 - 89) | 6<br>(3 - 9) | 22<br>(20 - 23) | 37<br>(31 - 42) | 38<br>(20 - 57) | 48<br>(43 - 54) | 293<br>(242 - 366) | 3441<br>(2091 - 7577) |
| 70 all | GSK | 965<br>(876 - 1064) | 82<br>(70 - 95) | 7<br>(4 - 10) | 23<br>(21 - 25) | 32<br>(27 - 37) | 33<br>(17 - 52) | 33<br>(29 - 38) | 248<br>(201 - 315) | 2794<br>(1639 - 6951) |
| | Pfizer | 1019<br>(925 - 1121) | 80<br>(68 - 93) | 7<br>(4 - 10) | 19<br>(17 - 20) | 33<br>(28 - 39) | 35<br>(18 - 54) | 41<br>(36 - 47) | 237<br>(193 - 301) | 2776<br>(1625 - 6923) |
| 75 all | GSK | 1024<br>(931 - 1128) | 90<br>(77 - 104) | 7<br>(4 - 11) | 18<br>(16 - 20) | 25<br>(21 - 30) | 29<br>(12 - 47) | 27<br>(23 - 30) | 199<br>(157 - 259) | 2197<br>(1177 - 5883) |
| | Pfizer | 1067<br>(970 - 1173) | 89<br>(76 - 102) | 7<br>(4 - 10) | 15<br>(13 - 16) | 26<br>(22 - 31) | 30<br>(13 - 48) | 33<br>(29 - 38) | 190<br>(151 - 247) | 1979<br>(1169 - 5851) |
| 80 all | GSK | 1071<br>(973 - 1179) | 97<br>(82 - 111) | 8<br>(4 - 11) | 14<br>(13 - 16) | 20<br>(15 - 25) | 23<br>(8 - 40) | 19<br>(16 - 22) | 139<br>(106 - 188) | 1597<br>(796 - 4890) |
| | Pfizer | 1105<br>(1004 - 1216) | 96<br>(81 - 110) | 8<br>(4 - 11) | 12<br>(10 - 13) | 21<br>(16 - 26) | 23<br>(9 - 41) | 23<br>(20 - 27) | 133<br>(102 - 180) | 1402<br>(752 - 4870) |
| 60 AR & 50 HR | GSK | 862<br>(784 - 948) | 72<br>(61 - 85) | 6<br>(3 - 9) | 31<br>(30 - 33) | 40<br>(35 - 45) | 41<br>(23 - 59) | 48<br>(43 - 54) | 388<br>(323 - 478) | 4571<br>(2870 - 9171) |
| | Pfizer | 935<br>(850 - 1028) | 70<br>(59 - 82) | 6<br>(3 - 9) | 25<br>(24 - 27) | 42<br>(37 - 47) | 42<br>(24 - 61) | 59<br>(53 - 66) | 372<br>(311 - 457) | 4552<br>(2733 - 9134) |
| | GSK | 877 | 72 | 6 | 30 | 40 | 41 | 47 | 364 | 4285 |

| | | | | | | | | | | |
|---|---|---|---|---|---|---|---|---|---|---|
| 65 AR & 50 HR | | (797 - 965) | (61 - 85) | (3 - 9) | (28 - 32) | (35 - 45) | (22 - 59) | (42 - 52) | (303 - 448) | (2689 - 8601) |
| | Pfizer | 947<br>(861 - 1041) | 70<br>(59 - 82) | 6<br>(3 - 9) | 24<br>(23 - 26) | 42<br>(37 - 47) | 42<br>(24 - 61) | 58<br>(52 - 65) | 349<br>(292 - 429) | 4265<br>(2561 - 8560) |
| 65 AR & 60 HR | GSK | 891<br>(810 - 980) | 75<br>(63 - 87) | 6<br>(3 - 9) | 29<br>(27 - 31) | 38<br>(33 - 43) | 39<br>(21 - 58) | 42<br>(38 - 47) | 327<br>(271 - 405) | 4002<br>(2323 - 8816) |
| | Pfizer | 958<br>(872 - 1054) | 73 (61 - 85) | 6<br>(3 - 9) | 23<br>(22 - 25) | 40<br>(35 - 45) | 41<br>(23 - 59) | 52<br>(46 - 58) | 314<br>(261 - 388) | 3686<br>(2310 - 8773) |
| 70 AR & 50 HR | GSK | 893<br>(812 - 983) | 73<br>(61 - 85) | 6<br>(3 - 9) | 29<br>(27 - 30) | 40<br>(35 - 45) | 41<br>(22 - 59) | 45<br>(41 - 51) | 337<br>(281 - 415) | 3965<br>(2488 - 7961) |
| | Pfizer | 960<br>(873 - 1056) | 70<br>(59 - 82) | 6<br>(3 - 9) | 23<br>(22 - 25) | 42<br>(37 - 47) | 42<br>(24 - 61) | 56<br>(50 - 63) | 323<br>(271 - 398) | 3945<br>(2369 - 7920) |
| 70 AR & 60 HR | GSK | 907<br>(825 - 998) | 75<br>(63 - 87) | 6<br>(3 - 9) | 28<br>(26 - 29) | 38<br>(33 - 43) | 39<br>(21 - 58) | 40<br>(36 - 45) | 300<br>(248 - 371) | 3654<br>(2122 - 8047) |
| | Pfizer | 972<br>(883 - 1068) | 73<br>(61 - 85) | 6<br>(3 - 9) | 22<br>(21 - 24) | 40<br>(35 - 45) | 41<br>(23 - 59) | 49<br>(44 - 56) | 287<br>(239 - 355) | 3367<br>(2109 - 8010) |
| 75 AR & 50 HR | GSK | 914<br>(832 - 1006) | 73<br>(61 - 85) | 6<br>(3 - 9) | 27<br>(25 - 29) | 40<br>(35 - 45) | 40<br>(22 - 59) | 44<br>(40 - 50) | 311<br>(259 - 383) | 3648<br>(2287 - 8649) |
| | Pfizer | 977<br>(889 - 1075) | 70<br>(59 - 83) | 6<br>(3 - 9) | 22<br>(20 - 23) | 42<br>(36 - 47) | 42<br>(24 - 61) | 55<br>(49 - 61) | 298<br>(250 - 367) | 3628<br>(2179 - 7287) |
| 75 AR & 60 HR | GSK | 928<br>(844 - 1022) | 75<br>(63 - 87) | 6<br>(3 - 9) | 26<br>(24 - 28) | 38<br>(33 - 43) | 39<br>(21 - 58) | 39<br>(35 - 43) | 272<br>(225 - 337) | 3308<br>(1921 - 7286) |
| | Pfizer | 989<br>(899 - 1087) | 73<br>(61 - 85) | 6<br>(3 - 9) | 21<br>(20 - 23) | 40<br>(35 - 45) | 41<br>(23 - 59) | 48<br>(43 - 54) | 261<br>(217 - 322) | 3050<br>(1910 - 7253) |
| 80 AR & 50 HR | GSK | 931<br>(847 - 1024) | 73<br>(61 - 85) | 6<br>(3 - 9) | 26<br>(24 - 27) | 40<br>(35 - 45) | 40<br>(22 - 59) | 43<br>(39 - 49) | 290<br>(242 - 358) | 3402<br>(2133 - 8078) |
| | Pfizer | 991 | 71 | 6 | 21 | 41 | 42 | 54 | 279 | 3383 |

| | | | | | | | | | | |
|---|---|---|---|---|---|---|---|---|---|---|
| | | (902 - 1089) | (59 - 83) | (3 - 9) | (19 - 22) | (36 - 47) | (24 - 61) | (48 - 60) | (233 - 342) | (2032 - 6796) |
| 80 AR & 60 HR | GSK | 945<br>(859 - 1040) | 75<br>(63 - 87) | 6<br>(3 - 9) | 25<br>(23 - 26) | 38<br>(33 - 43) | 39<br>(21 - 57) | 38<br>(34 - 42) | 250<br>(207 - 310) | 3040<br>(1767 - 6695) |
| | Pfizer | 1002<br>(912 - 1102) | 73<br>(61 - 85) | 6<br>(3 - 9) | 20<br>(18 - 21) | 40<br>(35 - 45) | 41<br>(22 - 59) | 46<br>(41 - 52) | 240<br>(200 - 297) | 2804<br>(1756 - 6670) |

*HR = high risk (one or more chronic medical condition); AR = average risk (no chronic medical conditions)

**Supplementary Table 3.** Costs, quality-adjusted life years, and incremental cost-effectiveness ratios for all vaccination strategies with vaccine effectiveness estimates based on (A) Arexvy (GSK) and (B) Abrysvo (Pfizer), for the societal perspective (human capital approach).

(A)

| Strategy* | Costs ($) | Effect (QALYs lost) | Incremental Costs ($) | Incremental Effect (QALYs gained) | Sequential ICER ($ per QALY) |
|---|---|---|---|---|---|
| 80 HR | 16,946,814 | 251.7969 | -- | -- | -- |
| 75 HR | 17,569,239 | 235.0853 | 622,425 | 16.7117 | 37,245 |
| 70 HR | 18,360,952 | 216.7820 | 791,713 | 18.3033 | 43,255 |
| 60 HR | 20,819,218 | 192.2692 | 2,458,266 | 24.5127 | 100,285 |
| 80 AR & 50 HR | 23,069,033 | 181.5907 | 2,249,815 | 10.6786 | 210,685 |
| 75 AR & 50 HR | 23,822,246 | 181.0419 | 753,213 | 0.5487 | 1,372,607 |
| 70 AR & 50 HR | 24,796,550 | 180.3799 | 974,304 | 0.6620 | 1,471,710 |
| 60 AR & 50 HR | 26,776,473 | 179.4767 | 1,979,922 | 0.9031 | 2,192,259 |
| 80 all | 17,456,862 | 250.3266 | -- | -- | Extended dominated |
| 65 HR | 19,547,848 | 205.8000 | -- | -- | Extended dominated |
| 80 AR & 60 HR | 21,329,266 | 190.7989 | -- | -- | Extended dominated |
| 75 AR & 60 HR | 22,082,479 | 190.2501 | -- | -- | Extended dominated |
| 70 AR & 60 HR | 23,056,783 | 189.5881 | -- | -- | Extended dominated |
| 65 AR & 50 HR | 25,801,755 | 179.9373 | -- | -- | Extended dominated |
| No vaccination | 17,773,068 | 293.0558 | -- | -- | Dominated |
| 75 all | 18,832,500 | 233.0662 | -- | -- | Dominated |
| 70 all | 20,598,516 | 214.1009 | -- | -- | Dominated |
| 65 all | 22,790,617 | 202.6763 | -- | -- | Dominated |
| 65 AR & 60 HR | 24,061,988 | 189.1455 | -- | -- | Dominated |
| 60 all | 25,036,705 | 188.6850 | -- | -- | Dominated |

(B)

| Strategy* | Costs ($) | Effect (QALYs lost) | Incremental Costs ($) | Incremental Effect (QALYs gained) | Sequential ICER ($ per QALY) |
|---|---|---|---|---|---|
| 80 HR | 16,919,230 | 250.5372 | -- | -- | -- |
| 75 HR | 17,530,551 | 233.2090 | 611,321 | 17.3283 | 35,279 |
| 70 HR | 18,316,097 | 214.3400 | 785,546 | 18.8690 | 41,632 |
| 60 HR | 20,779,007 | 189.2850 | 2,462,909 | 25.0550 | 98,300 |
| 80 AR & 50 HR | 23,049,217 | 178.4421 | 2,270,210 | 10.8429 | 209,373 |
| 75 AR & 50 HR | 23,811,507 | 177.9290 | 762,290 | 0.5131 | 1,485,658 |
| 70 AR & 50 HR | 24,799,954 | 177.3123 | 988,447 | 0.6167 | 1,602,893 |
| 60 AR & 50 HR | 26,800,376 | 176.4777 | 2,000,422 | 0.8346 | 2,396,720 |
| 80 all | 17,452,838 | 249.1642 | -- | -- | Extended dominated |
| 65 HR | 19,507,024 | 203.1625 | -- | -- | Extended dominated |
| 80 AR & 60 HR | 21,312,615 | 187.9120 | -- | -- | Extended dominated |
| 75 AR & 60 HR | 22,074,904 | 187.3989 | -- | -- | Extended dominated |
| 65 AR & 50 HR | 25,816,120 | 176.9007 | -- | -- | Extended dominated |
| No vaccination | 17,773,068 | 293.0558 | -- | -- | Dominated |
| 75 all | 18,826,449 | 231.3228 | -- | -- | Dominated |
| 70 all | 20,600,442 | 211.8372 | -- | -- | Dominated |
| 65 all | 22,807,534 | 200.2481 | -- | -- | Dominated |
| 70 AR & 60 HR | 23,063,352 | 186.7822 | -- | -- | Dominated |
| 65 AR & 60 HR | 24,079,517 | 186.3706 | -- | -- | Dominated |
| 60 all | 25,063,774 | 185.9475 | -- | -- | Dominated |

*HR = high risk (one or more chronic medical conditions); AR = average risk (no chronic medical conditions)

**Supplementary Table 4.** Costs, quality-adjusted life years, and incremental cost-effectiveness ratios for all vaccination strategies with vaccine effectiveness estimates based on (A) Arexvy (GSK) and (B) Abrysvo (Pfizer), for the societal perspective (friction cost approach).

(A)

| Strategy* | Costs ($) | Effect (QALYs lost) | Incremental Costs ($) | Incremental Effect (QALYs gained) | Sequential ICER ($ per QALY) |
|---|---|---|---|---|---|
| 80 HR | 15,276,101 | 251.7969 | -- | -- | -- |
| 75 HR | 15,940,028 | 235.0853 | 663,927 | 16.7117 | 39,728 |
| 70 HR | 16,798,771 | 216.7820 | 858,743 | 18.3033 | 46,917 |
| 60 HR | 19,451,470 | 192.2692 | 2,652,699 | 24.5127 | 108,217 |
| 80 AR & 50 HR | 21,917,284 | 181.5907 | 2,465,814 | 10.6786 | 230,913 |
| 75 AR & 50 HR | 22,671,217 | 181.0419 | 753,933 | 0.5487 | 1,373,920 |
| 70 AR & 50 HR | 23,646,744 | 180.3799 | 975,527 | 0.6620 | 1,473,557 |
| 60 AR & 50 HR | 25,629,981 | 179.4767 | 1,983,236 | 0.9031 | 2,195,929 |
| 80 all | 15,788,058 | 250.3266 | -- | -- | Extended dominated |
| 65 HR | 18,041,213 | 205.8000 | -- | -- | Extended dominated |
| 80 AR & 60 HR | 19,963,427 | 190.7989 | -- | -- | Extended dominated |
| 75 AR & 60 HR | 20,717,360 | 190.2501 | -- | -- | Extended dominated |
| 70 AR & 60 HR | 21,692,887 | 189.5881 | -- | -- | Extended dominated |
| 65 AR & 50 HR | 24,652,905 | 179.9373 | -- | -- | Extended dominated |
| No vaccination | 16,002,334 | 293.0558 | -- | -- | Dominated |
| 75 all | 17,205,918 | 233.0662 | -- | -- | Dominated |
| 70 all | 19,040,188 | 214.1009 | -- | -- | Dominated |
| 65 all | 21,288,790 | 202.6763 | -- | -- | Dominated |
| 65 AR & 60 HR | 22,699,047 | 189.1455 | -- | -- | Dominated |
| 60 all | 23,676,123 | 188.6850 | -- | -- | Dominated |

(B)

| Strategy* | Costs ($) | Effect (QALYs lost) | Incremental Costs ($) | Incremental Effect (QALYs gained) | Sequential ICER ($ per QALY) |
|---|---|---|---|---|---|
| 80 HR | 15,252,775 | 250.5372 | -- | -- | -- |
| 75 HR | 15,907,407 | 233.2090 | 654,632 | 17.3283 | 37,778 |
| 70 HR | 16,762,563 | 214.3400 | 855,156 | 18.8690 | 45,321 |
| 60 HR | 19,425,978 | 189.2850 | 2,663,415 | 25.0550 | 106,303 |
| 80 AR & 50 HR | 21,919,524 | 178.4421 | 2,493,546 | 10.8429 | 229,971 |
| 75 AR & 50 HR | 22,682,571 | 177.9290 | 763,047 | 0.5131 | 1,487,134 |
| 70 AR & 50 HR | 23,672,313 | 177.3123 | 989,742 | 0.6167 | 1,604,991 |
| 60 AR & 50 HR | 25,676,226 | 176.4777 | 2,003,913 | 0.8346 | 2,400,903 |
| 80 all | 15,788,390 | 249.1642 | -- | -- | Extended dominated |
| 65 HR | 18,010,516 | 203.1625 | -- | -- | Extended dominated |
| 80 AR & 60 HR | 19,961,594 | 187.9120 | -- | -- | Extended dominated |
| 75 AR & 60 HR | 20,724,641 | 187.3989 | -- | -- | Extended dominated |
| 70 AR & 60 HR | 21,714,382 | 186.7822 | -- | -- | Extended dominated |
| 65 AR & 50 HR | 24,689,532 | 176.9007 | -- | -- | Extended dominated |
| No vaccination | 16,002,334 | 293.0558 | -- | -- | Dominated |
| 75 all | 17,206,070 | 231.3228 | -- | -- | Dominated |
| 70 all | 19,050,967 | 211.8372 | -- | -- | Dominated |
| 65 all | 21,316,139 | 200.2481 | -- | -- | Dominated |
| 65 AR & 60 HR | 22,731,601 | 186.3706 | -- | -- | Dominated |
| 60 all | 23,718,295 | 185.9475 | -- | -- | Dominated |

*HR = high risk (one or more chronic medical conditions); AR = average risk (no chronic medical conditions)

**Supplementary Table 5.** Costs, quality-adjusted life years, and incremental cost-effectiveness ratios for age-based vaccination strategies only with vaccine effectiveness estimates based on (A) Arexvy (GSK) and (B) Abrysvo (Pfizer), for the health system perspective.

(A)

| Strategy | Costs ($) | Effect (QALYs lost) | Incremental Costs ($) | Incremental Effect (QALYs gained) | Sequential ICER ($ per QALY) |
|---|---|---|---|---|---|
| No vaccination | 11,462,844 | 293.0558 | -- | -- | -- |
| 80 all | 11,691,842 | 250.3266 | 228,998 | 42.7292 | 5,359 |
| 75 all | 13,140,984 | 233.0662 | 1,449,142 | 17.2604 | 83,958 |
| 70 all | 15,018,600 | 214.1009 | 1,877,616 | 18.9653 | 99,002 |
| 60 all | 19,407,496 | 188.6850 | 4,388,896 | 25.4159 | 172,683 |
| 65 all | 17,243,412 | 202.6763 | -- | -- | Extended dominated |

(B)

| Strategy | Costs ($) | Effect (QALYs lost) | Incremental Costs ($) | Incremental Effect (QALYs gained) | Sequential ICER ($ per QALY) |
|---|---|---|---|---|---|
| No vaccination | 11,462,844 | 293.0558 | -- | -- | -- |
| 80 all | 11,603,970 | 249.1642 | 141,126 | 43.8915 | 3,215 |
| 75 all | 13,029,887 | 231.3228 | 1,425,917 | 17.8414 | 79,922 |
| 70 all | 14,881,277 | 211.8372 | 1,851,390 | 19.4856 | 95,013 |
| 60 all | 19,247,044 | 185.9475 | 4,365,767 | 25.8897 | 168,630 |
| 65 all | 17,094,090 | 200.2481 | -- | -- | Extended dominated |
